# Spike-antibody responses to ChAdOx1 and BNT162b2 vaccines by demographic and clinical factors (Virus Watch study)

**DOI:** 10.1101/2021.05.12.21257102

**Authors:** Madhumita Shrotri, Ellen Fragaszy, Cyril Geismar, Vincent Nguyen, Sarah Beale, Isobel Braithwaite, Thomas E Byrne, Wing Lam Erica Fong, Jana Kovar, Annalan M D Navaratnam, Parth Patel, Anna Aryee, Jamie Lopez Bernal, Anne M Johnson, Alison Rodger, Andrew C Hayward, Robert W Aldridge

## Abstract

**Background:** Vaccination constitutes the best long-term solution against Coronavirus Disease 2019 (COVID-19). Real-world immunogenicity data are sparse, particularly for ChAdOx1 and in populations with chronic conditions; and given the UK’s extended dosing interval, it is also important to understand antibody responses in SARS-CoV-2-naive individuals following a single dose.

**Methods:** Adults aged ≥18 years from households enrolled in Virus Watch, a prospective community cohort study in England and Wales, provided capillary blood samples and self-reported vaccination status. Primary outcome variables were quantitative Spike total antibody levels (U/ml) and seropositivity to Spike (≥0.8 U/ml), as per Roche’s Elecsys Anti-SARS-CoV-2 S assay. Samples seropositive for Nucleocapsid, and samples taken prior to vaccination, were excluded. Outcomes were analysed by days since vaccination, vaccine type (BNT162b2 and ChAdOx1), and a range of self-reported demographic and clinical factors.

**Results:** 8,837 vaccinated participants (median age 65 years [IQR: 58, 71]), contributed 17,160 samples (10,508 following ChAdOx1, 6,547 following BNT162b2). Seropositivity to Spike was 96.79% (95% CI 96.42, 97.12) from 28 days following a single dose, reaching 99.34% (98.91, 99.60) from 14 days after a second dose. Seropositivity rates, and Spike-antibody levels rose more quickly following the first dose of BNT162b2, however, were equivalent for both vaccines by 4 and 8 weeks, respectively. There was evidence for lower S-antibody levels with increasing age (p=0.0001). In partially vaccinated 65-79 year-olds, lower S-antibody levels were observed in men compared with women (26.50 vs 44.01 U/ml, p<0.0001), those with any chronic condition (33.8 vs 43.83 U/ml, p<0.0001), diabetes (22.46 vs 36.90 U/ml, p<0.0001), cardiovascular disease (32.9 vs 37.9 U/ml, p=0.0002), obesity (27.2 vs 37.42, p<0.0001), cancer diagnosis (31.39 vs 36.50 U/ml, p=0.0001), particularly those with haematological cancers (7.94 vs 32.50 U/ml, p<0.0001), and for those currently on statin therapy (30.03 vs 39.39, p<0.0001), or on any immunosuppressive therapy (28.7 vs 36.78 U/ml, p<0.0001), particularly those on oral steroids (16.8 vs 36.07, p<0.0001). Following a second dose, high S-antibody titres (≥250U/ml) were observed across all groups.

**Interpretation:** A single dose of either BNT162b2 or ChAdOx1 leads to high Spike seropositivity rates in SARS-CoV-2-naive individuals. Observed disparities in antibody levels by vaccine type, age, and comorbidities highlight the importance of ongoing non-pharmaceutical preventative measures for partially vaccinated adults, particularly those who are older and more clinically vulnerable; and high antibody levels across all groups following a second dose demonstrate the importance of complete vaccination. However, the relationship between Spike-antibody levels and protection against COVID-19, and thus the clinical significance of observed disparities, is not yet clear.

## Introduction

The ongoing Severe Acute Respiratory Syndrome Coronavirus 2 (SARS-CoV-2) pandemic and the morbidity and mortality resulting from the associated clinical syndrome, Coronavirus disease-2019 (COVID-19), has had a devastating impact on many nations with 163,312,429 confirmed cases and 3,386,825 deaths reported globally as of 18 May 2021^1^. Early in the pandemic, global vaccine developers joined the race to find a definitive, long-term solution, pivoting both established and experimental vaccine platforms towards SARS-CoV-2, with many candidates having now completed clinical testing and achieved licensure^2^. Pfizer/BioNTech’s BNT162b2 messenger RNA vaccine, and Oxford/AstraZeneca’s ChAdOx1 nCoV-19 non-replicating adenovirus-vectored vaccine were licensed in the UK in December 2020^3,4^. Both vaccines are based on the Spike protein of SARS-CoV-2, which contains the receptor binding domain - the target for neutralising antibodies that can prevent the virus binding to the ACE2-receptor on the human cell surface^5^. The formulation of these vaccines allows antibodies against the Nucleocapsid protein, an abundant and highly immunogenic viral antigen^6^, to remain discriminatory for natural infection. Vaccination has been offered in accordance with the UK’s Joint Committee on Vaccination and Immunisation’s prioritisation framework^7^, with 36,811,405 people having received their first vaccine dose as of 18 May 2021^8^.

Trial and observational data have demonstrated the efficacy of ChAdOx1 and BNT162b2 against infection and clinical severity^9–12^, with increasing evidence for impact on transmission^11,13–15^. While there are abundant trial data on immunogenicity, evidence is generally limited to younger, healthier trial populations, as well as to manufacturer-recommended dosing regimens. In the UK, the recommended dosing interval was extended from 3-4 weeks, to 8-12 weeks in order to maximise first-dose coverage across the population^16^; thus, there is a pressing need to understand features of the humoral immune response between 4-12 weeks in a real-world setting. Furthermore, it is critical to understand immunogenicity amongst older people, those with metabolic risk factors, and those from ethnic minority backgrounds, as these groups are at highest risk of COVID-19 morbidity and mortality^17–20^; as well as those on immunosuppressive therapies, which are likely to attenuate vaccine responses^21^. The existing observational data are limited by reporting on specific populations such as healthcare workers^22,23^ or long-term care residents^24^; by focusing solely on BNT162b2^22,25,26^ and with shorter dosing intervals^25^; by lack of detailed information on underlying health conditions^27^; and often relatively small sample sizes.

We analysed serological data from a large, prospective community cohort, Virus Watch^28^, obtained using a widely available, validated commercial assay. We determined seropositivity rates for Spike, and Spike-antibody levels in order to investigate vaccine responses in individuals without prior infection from the general population of England and Wales.

## Methods

### Study design and setting

This analysis was conducted as part of a prospective community cohort study based in England and Wales, which commenced recruitment in May 2020 (detailed methods are described elsewhere)^28^. Invitations to participate in monthly antibody testing were sent to previously enrolled eligible households over February-March 2021. Consenting participants provided monthly capillary blood samples between 24 February and 11 May 2021, and self-reported vaccination data from 11 January 2021, in addition to demographic and clinical data at enrollment.

Blood samples (400-600 microlitres) were self-collected by participants using an at-home capillary blood sample collection kit manufactured by Thriva Ltd [https://thriva.co/]. Completed kits were returned by participants using pre-paid envelopes and priority postage boxes to UKAS-accredited laboratories. The sampling date was taken as being two days prior to the date of receipt by the laboratory. Serological testing was undertaken using Roche’s Elecsys Anti-SARS-CoV-2 assays targeting total immunoglobulin (Ig) to the Nucleocapsid (N) protein or to the receptor binding domain in the S1 subunit of the Spike protein (S) (Roche Diagnostics, Basel, Switzerland). At the manufacturer-recommended cut-offs (≥0.1 cut-off index [COI] for N and ≥0.8 units per millilitre [U/ml] for S), the N assay has a sensitivity of 97·2%-99.5% and specificity of 99.8%^29–31^, while the S assay has a sensitivity of 97.9%-98.8% and a specificity of 100%^32–34^, with high agreement between assays for samples from previously infected individuals^32^.

### Participants

Within the Virus Watch cohort (Table S1), eligible households were defined as having at least one adult aged 18 years and over, a valid England or Wales postcode, a complete postal address registered at enrollment, complete gender and ethnicity information for all household members, and not enrolled in the study sub-cohort undergoing longitudinal point-of-care antibody testing as part of index case investigations^28^. Individuals aged ≥18 years within eligible households were provided with a participant information sheet and gave valid, informed consent through an electronic REDCap form.

Only samples taken on the same day as, or in the days following the first dose of vaccination were included in this analysis. Evidence of natural infection was defined as seropositivity for the Nucleocapsid protein - samples meeting this criterion were excluded from the analysis so as to investigate vaccine responses only, and not combined with infection responses. Samples with void results from either assay were excluded from the analysis. Samples recorded as being 12 or more weeks after the first vaccine dose were also excluded due to the high likelihood the participant had omitted reporting their second vaccine dose.

### Exposure variables

Self-reported vaccination status was collected through the weekly Virus Watch questionnaire. The item was introduced on 11 January 2021 and asked about any prior vaccination for the first two surveys. Subsequently, participants were asked to provide a weekly update only (Table S2). Self-reported vaccination data underwent cleaning and individuals with implausible vaccination dates, for example those reporting vaccination prior to national licensure dates or those reporting a dose interval of <21 days, and where the correct date could not be ascertained from other survey responses, were excluded from the analysis as these were assumed either to be clinical trial participants or to have reported an erroneous date. Individuals who reported vaccination but did not provide dates were also excluded.

Time since vaccination was defined according to whether a sample was assigned as being taken on the same date as, or subsequent to, the first vaccine dose, and prior to the date of the second dose (first dose sample); or on the same date as, or subsequent to, the second dose (second dose sample). Days since vaccination were grouped into 7- or 14-day intervals following the first and second doses with binary variables to indicate results that were ≥28 days following the first dose, and ≥14 days following the second dose.

### Outcome variables

The main outcome variables were seropositivity to Spike, and Spike antibody level (U/ml), as per Roche’s Elecsys Anti-SARS-CoV-2 S assay, in the absence of seropositivity to Nucleocapsid. The assay measuring range was 0.4□250 U/mL, with the threshold for seropositivity to Spike defined as ≥0.8 U/ml.

### Covariates

Self-reported age, sex, ethnicity, height, and weight, as well as binary clinical variables, including current statin or immunosuppressive therapy, cancer diagnosis (previous or current), chronic conditions, and receipt of a COVID-19 clinical risk letter from the NHS (a proxy for shielding status during the ‘first wave’), were collected at enrollment (Table S2). Where appropriate, individual conditions were grouped into broader categories such as respiratory, neurological, or cardiovascular conditions. Obesity was assigned by the research team as BMI ≥30, calculated using self-reported height and weight data, and was not included within self-reported ‘chronic condition’ status.

Age was grouped into 18-34, 35-49, 50-64, 65-79, ≥80 years categories. Ethnicity data were collapsed into White, South Asian, Other Asian, and Mixed categories. Sex was limited to Male and Female with other categories suppressed due to small numbers. Vaccine types other than BNT162b2 and ChAdOx1 were not examined separately due to small numbers.

### Statistical analysis

We calculated proportions of seropositive individuals (as per ≥0.8 U/ml cut-off) for each time interval and the 95% confidence interval around each proportion. We calculated the median and interquartile range of S-antibody levels for each group from all available results, including samples below the cut-off for binary seropositivity.

The distribution of S-antibody levels was tested for normality using the Kolmogorov-Smirnov test. Bonferroni-adjusted p-values (threshold for evidence set at p<0.0017) for the difference between median S-antibody levels between different groups were derived using non-parametric tests: the Mann-Whitney U (Wilcoxon Rank Sum) test for two groups, or the Kruskal-Wallis test for >2 groups. We compared S-antibody levels between age groups, at ≥28 days after the first dose. We also compared S-antibody levels by sex, ethnicity, and chronic condition, obesity, or immunosuppressive therapy status at ≥28 days after the first dose. To minimise confounding by age this analysis was restricted to 65-79 year-olds for most conditions as this was the largest age-group; however for HIV the 35-49 years age-group was used as it contained most individuals with HIV. We also compared S-antibody levels between 65-79 year-old BNT162b2 and ChAdOx1 vaccinees at various time intervals after the first dose. We did not compare S-antibody levels after the second dose as most individuals had reached the assay limit of 250 U/ml.

Analysis was conducted in Stata version 16.0 (StataCorp, TX, USA) and R (version 4.0.3).

### Ethical approval

This study has been approved by the Hampstead NHS Health Research Authority Ethics Committee. Ethics approval number - 20/HRA/2320.

## Results

8,837 participants without evidence of prior infection, with a median age of 65 (IQR 58, 71), of whom 57% were female and 97% were of a White ethnicity, contributed 17,160 samples following at least one dose of a COVID-19 vaccine (Table 1). 69% (n=6,141) of individuals reported at least one chronic condition, with 8.9% (n=790) reporting having received a clinical risk letter from the NHS, 10% (n=893) reporting ever having received a cancer diagnosis and 0.9% (n=76) reporting a haematological cancer diagnosis (leukaemia or lymphoma). 21% (n=1,774) were classified as obese, 26% (n=2,338) reported statin therapy, and 16% (n=1,384) reported immunosuppressive therapy, including oral (n=169) or inhaled (n=911) steroids, non-steroid immunosuppressants (n=274), immunosuppressive cancer therapy (n=74), or medications following organ transplant (n=22). 3,151 BNT162b2 vaccinees contributed 6,547 samples; and 5,618 ChAdOx1 vaccinees contributed 10,508 samples (Table 2). Demographic and clinical characteristics were broadly similar across the two main vaccine types, though BNT162b2 vaccinees were slightly older and more likely to report a clinical risk factor, and also to have longer median intervals between the first or second dose and sample collection (first dose: 47 vs 38 days; second dose: 20 vs 12 days).

**Table 1.** Demographic and clinical characteristics of included individuals and samples.

| **Table 1** | **Individuals** | **Samples** |
| --- | --- | --- |
|  | *N (%)* | *N (%)* |
| **Total** | 8,837 | 17,160 |
| Female sex | 5,045 (57%) | 9,879 (58%) |
| Age [years] - median (IQR) | 65 (58, 71) | 66 (60, 72) |
| **Age group** |  |  |
| 18-34 | 196 (2.2%) | 308 (1.8%) |
| 35-49 | 772 (8.7%) | 1,174 (6.8%) |
| 50-64 | 3,193 (36%) | 5,644 (33%) |
| 65-79 | 4,456 (50%) | 9,565 (56%) |
| 80+ | 219 (2.5%) | 468 (2.7%) |
| **Ethnicity** |  |  |
| White | 8,593 (97%) | 16,737 (98%) |
| Black | 21 (0.2%) | 38 (0.2%) |
| South Asian | 94 (1.1%) | 162 (0.9%) |
| Other Asian | 44 (0.5%) | 74 (0.4%) |
| Mixed | 47 (0.5%) | 82 (0.5%) |
| Other/Missing | 37 (0.4%) | 66 (0.4%) |
| **Clinical factors** |  |  |
| NHS Risk letter | 790 (8.9%) | 1,601 (9.3%) |
| Cancer diagnosis (previous or current) | 893 (10%) | 1,887 (11%) |
| - Haematological | 76 (0.9%) | 170 (1.0%) |
| - Non-haematological (solid-organ) | 825 (9.3%) | 1,738 (10%) |
| - Bowel | 89 (10.0%) | 193 (10%) |
| - Breast | 282 (32%) | 581 (31%) |
| - Liver | 6 (0.7%) | 13 (0.7%) |
| - Lung | 15 (1.7%) | 29 (1.5%) |
| - Prostate | 149 (17%) | 319 (17%) |
| - Skin | 164 (18%) | 354 (19%) |
| - Other | 183 (20%) | 381 (20%) |
| Obesity | 1,774 (21%) | 3,377 (21%) |
| No chronic conditions | 2,696 (31%) | 4,930 (29%) |
| HIV | 15 (0.2%) | 30 (0.2%) |
| Respiratory condition | 1,588 (18%) | 3,061 (18%) |
| Diabetes | 713 (8.1%) | 1,429 (8.3%) |
| Cardiovascular disease | 2,961 (34%) | 6,059 (35%) |
| Hypothyroidism | 668 (7.6%) | 1,344 (7.8%) |
| Chronic kidney disease | 135 (1.5%) | 274 (1.6%) |
| Liver disease | 178 (2.0%) | 344 (2.0%) |
| Neurological condition | 206 (2.3%) | 403 (2.3%) |
| Statin therapy | 2,338 (26%) | 4,872 (28%) |
| Immunosuppressive therapy - Any | 1,384 (16%) | 2,773 (16%) |
| - Steroid tablets | 169 (1.9%) | 347 (2.0%) |
| - Steroid inhaler | 911 (10%) | 1,787 (10%) |
| - Non-steroid immunosuppressive medication | 274 (3.1%) | 574 (3.3%) |
| - Immunosuppressive cancer therapy (chemotherapy, targeted therapy, radiotherapy) | 74 (0.8%) | 156 (0.9%) |
| - Immunosuppression following organ transplant | 22 (0.2%) | 41 (0.2%) |
| **Vaccination** |  |  |
| *Month of first vaccine dose* |  |  |
| December 2020 | 182 (2.1%) |  |
| January 2021 | 2351 (26.6%) |  |
| February 2021 | 4149 (47.0%) |  |
| March 2021 | 1859 (21.0%) |  |
| April 2021 | 162 (1.8%) |  |
| May 2021 | 14 (0.2%) |  |
| Missing date | 120 (1.36) |  |
| Received two vaccine doses | 3,395 (38.4%) |  |
| Dose interval [days] - median (IQR) | 77 (71, 78) |  |
| *Vaccine type* |  |  |
| BNT162b2 (Pfizer/BioNTech) | 3,151 (35.7%) |  |
| ChAdOx1 (Oxford/AstraZeneca) | 5,618 (63.6%) |  |
| mRNA-1273 (Moderna) | 18 (0.2%) |  |
| Other/Don't know/Missing | 50 (0.6%) |  |

**Table 2.** Key demographic and clinical features of individuals vaccinated with BNT162b2 and ChAdOx1, and the samples from these individuals.

| Table 2 | BNT162b2 (Pfizer/BioNTech) |  | ChAdOx1 (Oxford/AstraZeneca) |  |
| --- | --- | --- | --- | --- |
|  | Individuals | Samples | Individuals | Samples |
|  | *N (%)* | *N (%)* | *N (%)* | *N (%)* |
| Total | 3,151 | 6,547 | 5,618 | 10,508 |
| Female sex | 1,798 (57%) | 3,762 (57%) | 3,213 (57%) | 6,064 (58%) |
| Age [years] - median (IQR) | 68 (61, 73) | 68 (62, 73) | 64 (56, 69) | 65 (59, 70) |
| White ethnicity | 3,051 (97%) | 6,348 (97%) | 5,479 (98%) | 10,290 (98%) |
| NHS Risk Letter | 337 (11%) | 684 (10%) | 451 (8.0%) | 913 (8.7%) |
| Immunosuppressive medication (current) - Any | 549 (17%) | 1,144 (17%) | 827 (15%) | 1,611 (15%) |
| Cancer diagnosis (prior or current) - Any | 365 (12%) | 797 (12%) | 526 (9.4%) | 1,086 (10%) |
| No chronic conditions | 769 (24%) | 1,570 (24%) | 1,889 (34%) | 3,308 (31%) |
| Respiratory condition | 589 (19%) | 1,198 (18%) | 991 (18%) | 1,848 (18%) |
| Diabetes | 307 (9.7%) | 632 (9.7%) | 404 (7.2%) | 792 (7.5%) |
| Cardiovascular disease | 1,235 (39%) | 2,618 (40%) | 1,713 (30%) | 3,419 (33%) |
| Obesity | 608 (20%) | 1,235 (20%) | 1,151 (22%) | 2,114 (21%) |
| ***Vaccine timing*** |  |  |  |  |
| First dose sample |  | 4269 (65.2%) |  | 8778 (83.5%) |
| Days since first dose - median (IQR) |  | 47 (32, 62) |  | 38 (24, 55) |
| First dose 28+ days |  | 3592 (54.9%) |  | 6000 (57.1%) |
| Days since first dose if 28+ days - median (IQR) |  | 53 (40, 64) |  | 48 (37, 60) |
| Second dose sample |  | 2278 (34.8%) |  | 1730 (16.5%) |
| Days since second dose - median (IQR) |  | 20 (9, 34) |  | 12 (6, 21) |
| Second dose 14+ days |  | 1451 (22.2%) |  | 798 (7.6%) |
| Days since second dose if 14+ days - median (IQR) |  | 30 (20, 43) |  | 22 (17, 28) |

### Qualitative seropositivity to Spike

Seropositivity rates rose more quickly following the first dose of BNT162b2 than ChAdOx1 (Figure 1, Table 3); these were 92.24% (95% CI 88.8, 95.68) and 78.07% (75.14, 81.0) at 14-20 days, rising to 96.59% (94.77, 98.41) and 92.10% (90.47, 93.73) at 21-27 days, for BNT162b2 and ChAdOx1, respectively. By days 28-34, the proportions of seropositive individuals were similar for both vaccines (96.65% [95.17, 98.13] vs 96.0% [94.85, 97.15]), and remained around this level over 12 weeks following the first dose. Seropositivity rates were >99% for both vaccines from ≥14 days following the second dose.

**Table 3.**
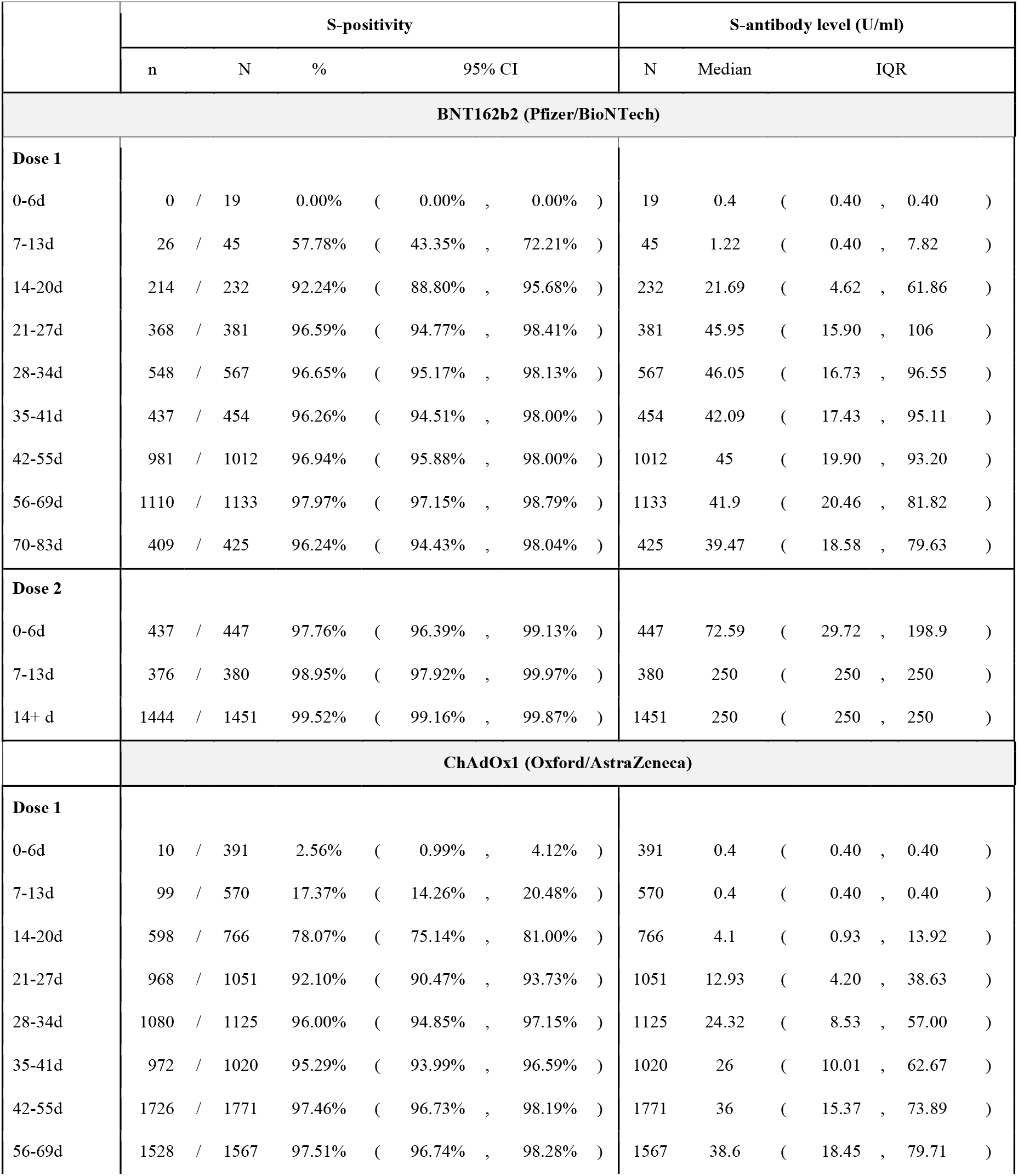
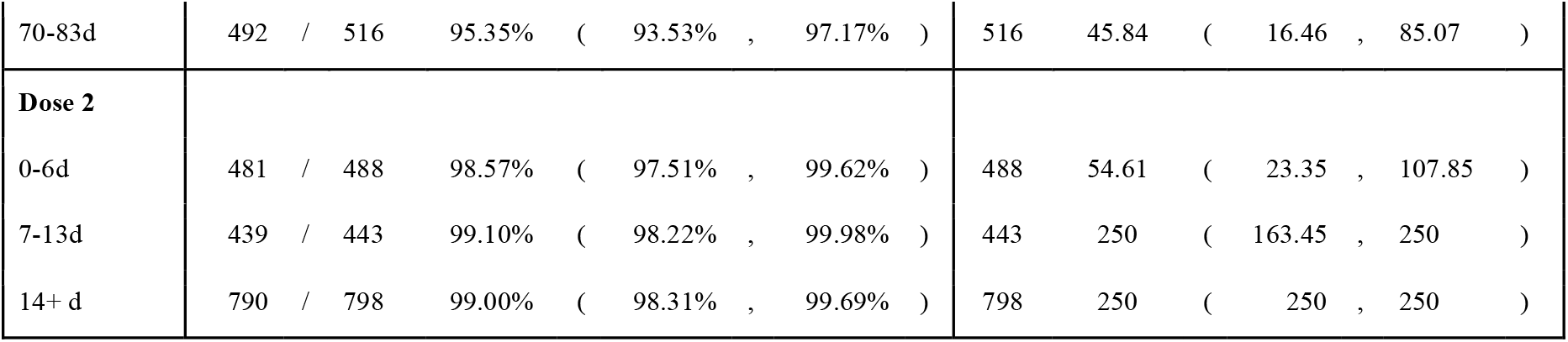
Data underlying Figure 1. Numbers and proportions of samples positive for S-antibodies (≥0.8 U/ml), and average S-antibody levels, by vaccine type and time since first or second dose of vaccination (all ages).

**Figure 1:**
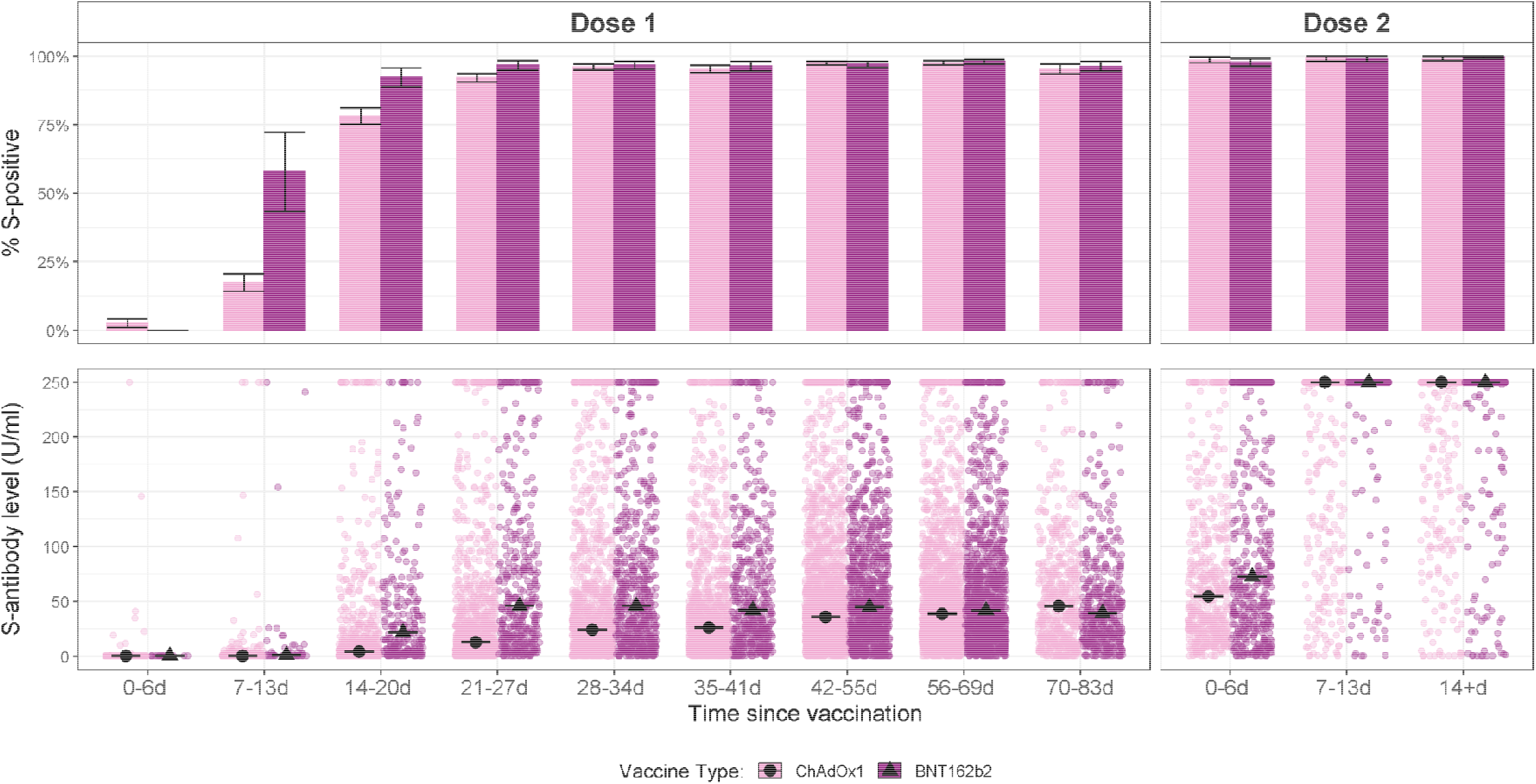
S-antibody positivity rates (as per ≥0.8 U/ml cutoff) and S-antibody levels (U/ml) by vaccine type and time since vaccination (all ages; black lines indicate median values).

At ≥28 days after one dose, a slightly higher proportion of females (ChAdOx1: 97.74% [97.19, 98.19] and BNT162b2: 97.68% [96.92, 98.25]) had seroconverted to Spike compared to males (ChAdOx1: 95.15% [94.25, 95.92] and BNT162b2: 96.23% [95.16, 97.07]), however there was no difference between groups at ≥14 days following a second dose of either vaccine (Table 4).

**Table 4.** Numbers and proportions of samples positive for S-antibodies (≥0.8 U/ml), and average S-antibody levels, at 28+ days since the first or 14+ days since the second dose of vaccination, by sex and vaccine type (all ages).

| Table 4 |  | S-positivity rate |  |  |  |  | S-antibody level (U/ml) |  |  |
| --- | --- | --- | --- | --- | --- | --- | --- | --- | --- |
| Sex | Vaccine type | n | / | N | % | 95% CI | N | Median | IQR |
| Dose 1 (28+ days) |  |  |  |  |  |  |  |  |  |
| Male | ChAdOx1 | 2415 | / | 2538 | 95.15% | ( 94.25% , 95.92% ) | 2538 | 24.15 | ( 9.70 , 56.58 ) |
|  | BNT162b2 | 1505 | / | 1564 | 96.23% | ( 95.16% , 97.07% ) | 1564 | 35.19 | ( 15.80 , 74.12 ) |
| Female | ChAdOx1 | 3377 | / | 3455 | 97.74% | ( 97.19% , 98.19% ) | 3455 | 39.4 | ( 17.97 , 84.94 ) |
|  | BNT162b2 | 1975 | / | 2022 | 97.68% | ( 96.92% , 98.25% ) | 2022 | 50.95 | ( 22.90 , 99.10 ) |
| Dose 2 (14+ days) |  |  |  |  |  |  |  |  |  |
| Male | ChAdOx1 | 342 | / | 347 | 98.56% | ( 96.58% , 99.40% ) | 347 | 250 | ( 250 , 250 ) |
|  | BNT162b2 | 598 | / | 601 | 99.50% | ( 98.46% , 99.84% ) | 601 | 250 | ( 250 , 250 ) |
| Female | ChAdOx1 | 448 | / | 451 | 99.33% | ( 97.95% , 99.79% ) | 451 | 250 | ( 250 , 250 ) |
|  | BNT162b2 | 843 | / | 847 | 99.53% | ( 98.75% , 99.82% ) | 847 | 250 | ( 250 , 250 ) |

Seropositivity rates were similar across all age groups for both vaccine types following first and second doses (Figure 2, Table 5). Amongst those aged ≥80 years, seropositivity rates were 83.87% (70.92, 96.82) and 95.1% (91.57, 98.64) after the first dose, and 100% and 99.07% (97.79, 100) after the second dose, of ChAdOx1 and BNT162b2 respectively.

**Table 5.** Data underlying Figure 2. Numbers and proportions of samples positive for S-antibodies (≥0.8 U/ml), and average S-antibody levels, at 28+ days since the first or 14+ days since the second dose of vaccination, by age group and vaccine type.

| Table 5 |  | S-positivity rate |  |  |  |  | S-antibody level (U/ml) |  |  |
| --- | --- | --- | --- | --- | --- | --- | --- | --- | --- |
| Age | Vaccine | n | / | N | % | 95% CI | N | Median | IQR |
| **Dose 1 (28+ days)** |  |  |  |  |  |  |  |  |  |
| **18-34y** | ChAdOx1 | 96 | / | 100 | 96.00% | ( 92.16% , 99.84% ) | 100 | 49.05 | ( 19.62 , 92.14 ) |
|  | BNT162b2 | 54 | / | 57 | 94.74% | ( 88.94% , 100% ) | 57 | 75.75 | ( 42.06 , 119.00 ) |
| **35-49y** | ChAdOx1 | 346 | / | 357 | 96.92% | ( 95.13% , 98.71% ) | 357 | 32.55 | ( 13.93 , 71.05 ) |
|  | BNT162b2 | 171 | / | 179 | 95.53% | ( 92.50% , 98.56% ) | 179 | 55.77 | ( 26.32 , 114.90 ) |
| **50-64y** | ChAdOx1 | 2231 | / | 2296 | 97.17% | ( 96.49% , 97.85% ) | 2296 | 32.995 | ( 14.49 , 74.36 ) |
|  | BNT162b2 | 794 | / | 814 | 97.54% | ( 96.48% , 98.61% ) | 814 | 48.105 | ( 22.12 , 101.45 ) |
| **65-79y** | ChAdOx1 | 3099 | / | 3215 | 96.39% | ( 95.75% , 97.04% ) | 3215 | 32.27 | ( 13.20 , 70.37 ) |
|  | BNT162b2 | 2329 | / | 2397 | 97.16% | ( 96.50% , 97.83% ) | 2397 | 40.6 | ( 17.80 , 82.20 ) |
| **80+y** | ChAdOx1 | 26 | / | 31 | 83.87% | ( 70.92% , 96.82% ) | 31 | 11.9 | ( 2.55 , 48.27 ) |
|  | BNT162b2 | 136 | / | 143 | 95.10% | ( 91.57% , 98.64% ) | 143 | 30.4 | ( 9.62 , 72.72 ) |
| **Dose 2 (14+ days)** |  |  |  |  |  |  |  |  |  |
| **18-34y** | ChAdOx1 | 11 | / | 11 | 100% | ( 100% , 100% ) | 11 | 250 | ( 123.9 , 250 ) |
|  | BNT162b2 | 34 | / | 34 | 100% | ( 100% , 100% ) | 34 | 250 | ( 250 , 250 ) |
| **35-49y** | ChAdOx1 | 15 | / | 15 | 100% | ( 100% , 100% ) | 15 | 250 | ( 250 , 250 ) |
|  | BNT162b2 | 108 | / | 109 | 99.08% | ( 97.29% , 100% ) | 109 | 250 | ( 250 , 250 ) |
| **50-64y** | ChAdOx1 | 91 | / | 92 | 98.91% | ( 96.79% , 100% ) | 92 | 250 | ( 250 , 250 ) |
|  | BNT162b2 | 312 | / | 315 | 99.05% | ( 97.98% , 100% ) | 315 | 250 | ( 250 , 250 ) |
| **65-79y** | ChAdOx1 | 661 | / | 668 | 98.95% | ( 98.18% , 99.72% ) | 668 | 250 | ( 250 , 250 ) |
|  | BNT162b2 | 777 | / | 778 | 99.87% | ( 99.62% , 100% ) | 778 | 250 | ( 250 , 250 ) |
| **80+y** | ChAdOx1 | 12 | / | 12 | 100% | ( 100% , 100% ) | 12 | 250 | ( 250 , 250 ) |
|  | BNT162b2 | 213 | / | 215 | 99.07% | ( 97.79% , 100% ) | 215 | 250 | ( 250 , 250 ) |

**Figure 2:**
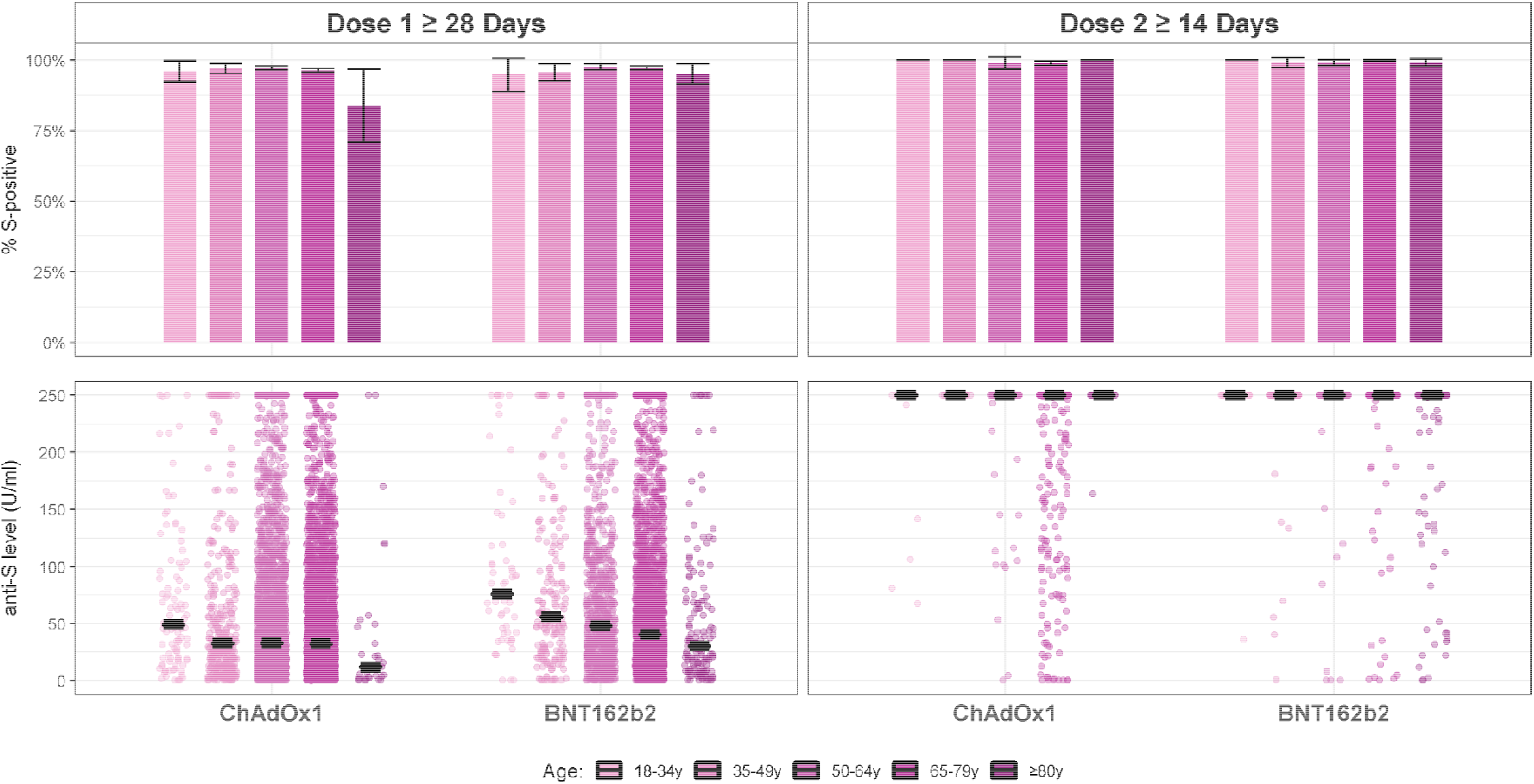
S-antibody positivity rates (as per ≥0.8 U/ml cutoff) and S-antibody levels (U/ml) by age group and vaccine type at 28+ days following the first dose and 14+ days following the second dose (black lines indicate median values).

At ≥28 days after the first dose, high seropositivity rates were observed amongst those reporting no chronic conditions (97.86% [97.31, 98.4]) (Figure 3, Table 6). However, seropositivity was as low as 72.28% (63.55, 81.01) amongst individuals with a history of haematological malignancy; 85.81% (80.31, 91.30) amongst those with chronic kidney disease (CKD); and 89.01% (87.02, 90.99) amongst those who received an NHS risk letter. Lower seropositivity rates were also seen amongst those on immunosuppressive therapies, with seropositivity of 32.0% (13.71, 50.29) amongst organ transplant recipients; 82.61% (74.86, 90.35) amongst cancer therapy recipients; 81.0% (75.56, 86.44) amongst those on oral steroid therapy; and 87.8% (84.26, 91.35) of those on non-steroidal immunosuppressants, at ≥28 days after a single dose. However, seropositivity rates were high amongst those using inhaled steroids (97.11% [96.07, 98.15]). At ≥14 days following the second dose, disparities between clinical groups had reduced substantially: seropositivity rates were 88.57% (78.03, 99.11) for haematological malignancy; 89.58% (80.94, 98.23) for CKD; 97.19% (95.28, 99.11) for NHS risk letters; 85.71 (59.79, 100%) for immunosuppression following organ transplant; 92.86% (83.32, 100%) for cancer therapy; 93.85% (88.0, 99.69) for oral steroids; and 98.85% (96.61, 100%) for non-steroidal immunosuppressants.

**Table 6.**
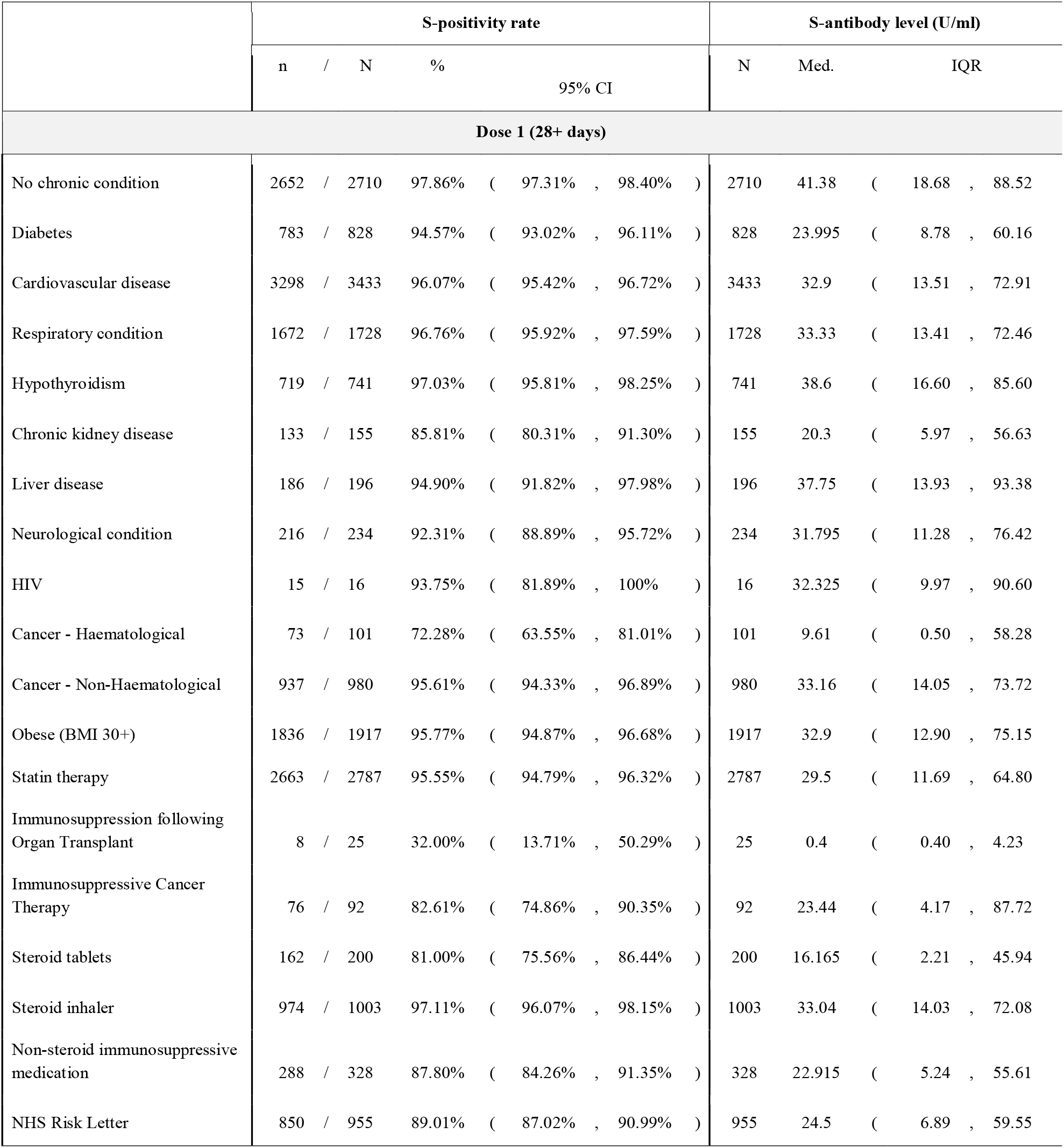
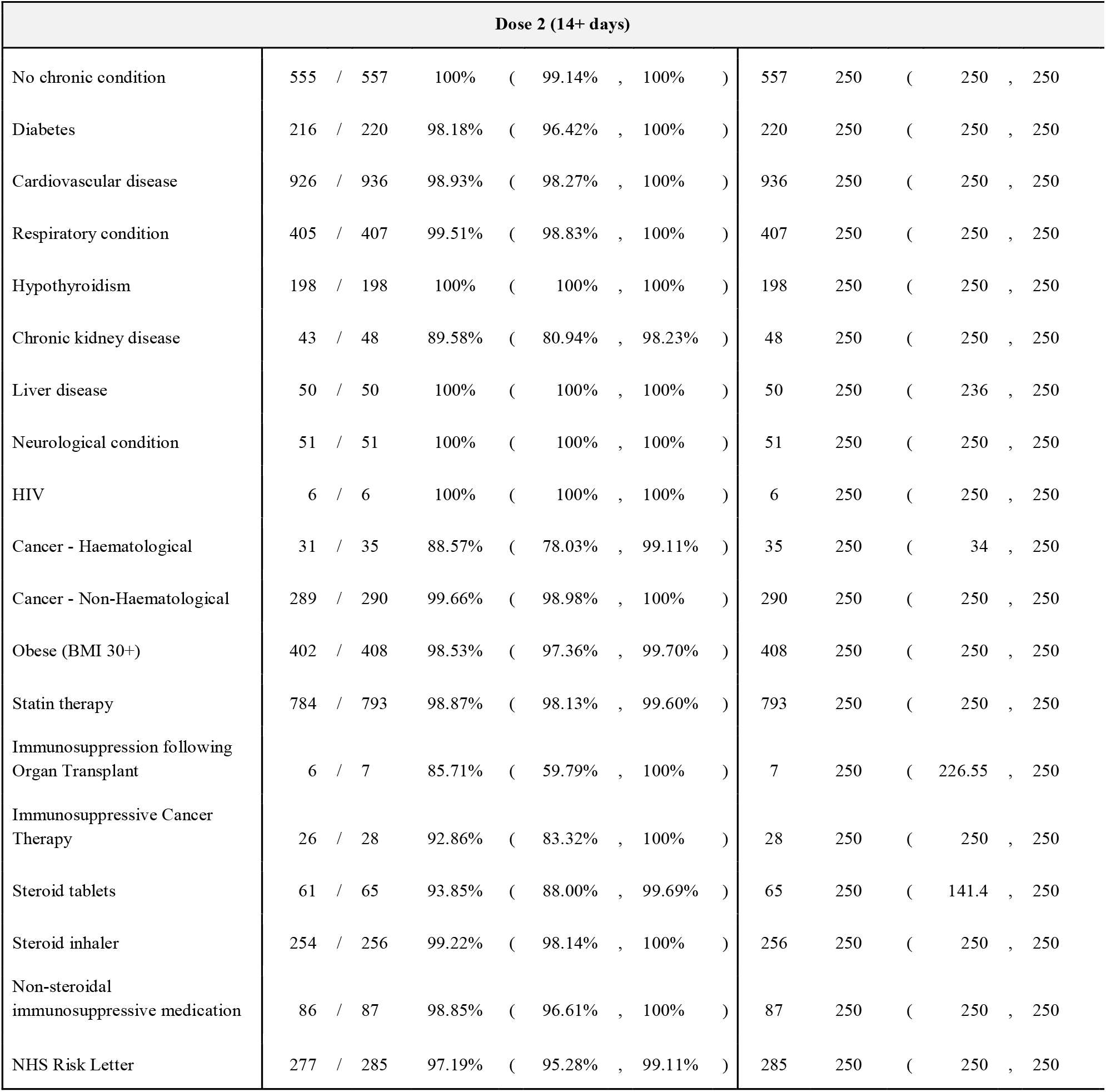
Data underlying Figure 3. Numbers and proportions of samples positive for S-antibodies (≥0.8 U/ml), and average S-antibody levels, at 28+ days since the first or 14+ days since the second dose of vaccination, by clinical variables (all ages).

**Figure 3:**
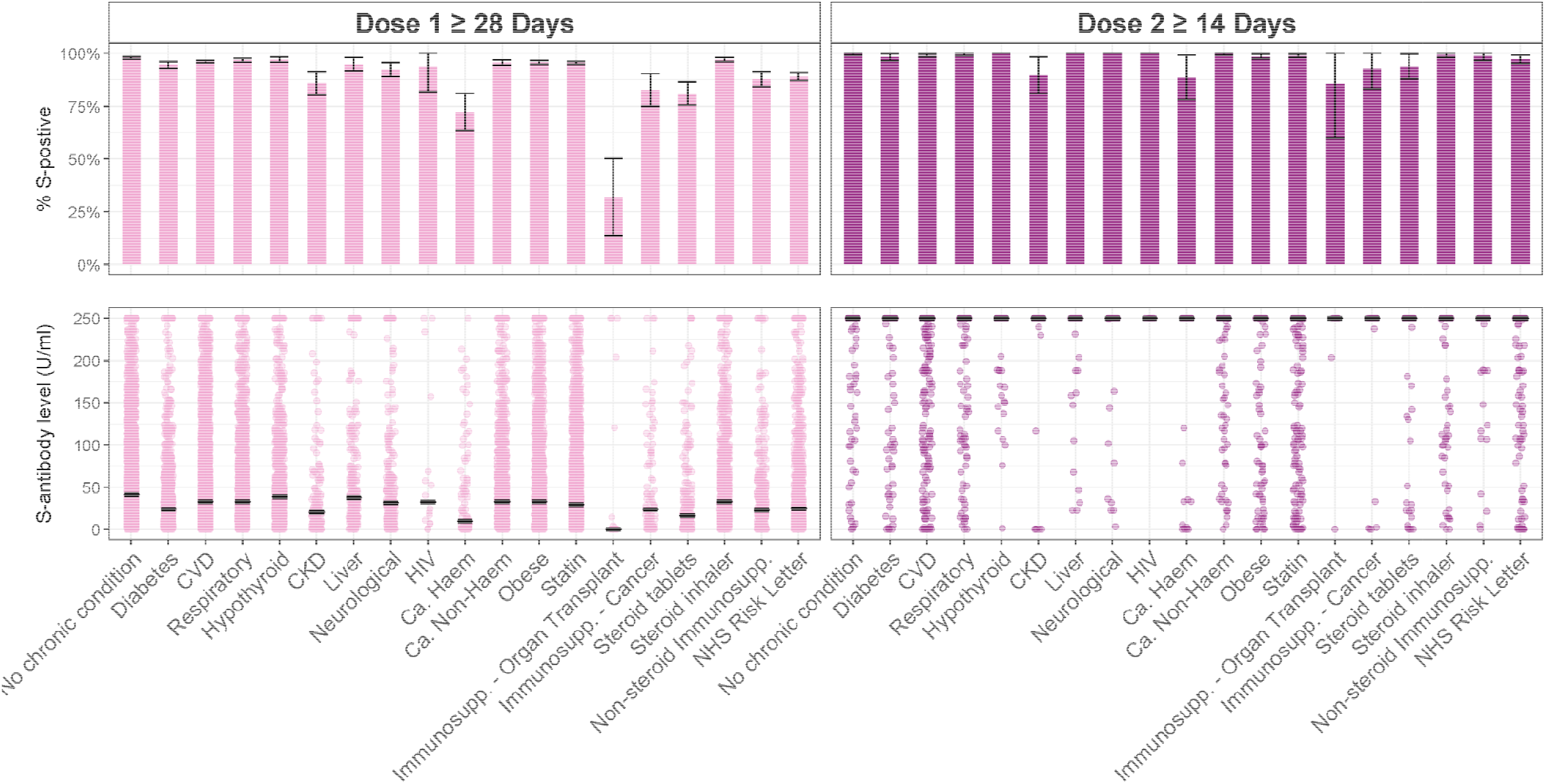
S-positivity rates (as per ≥0.8 U/ml cutoff) and S-antibody levels (U/ml) by a variety of clinical factors at 28+ days following the first dose and 14+ days following the second dose (all ages; black lines indicate median values).

### Quantitative Spike-antibody levels

Spike (S)-antibody levels increased over time after the first vaccine dose for both main vaccine types, however levels appeared higher at earlier time points following BNT162b2 than ChAdOx1 (Figure 1, Table 3). Amongst 65-79 year-olds, there was evidence for a true difference in antibody levels between the two vaccine types at 21-27 (BNT162b2: 37.71 U/ml vs ChAdOx1: 13.7 U/ml; p<0.0001), 28-34 (44.9 vs 21.2; p<0.0001), 35-41 (41.95 vs 23.4; p<0.0001), and 42-55 days (42.39 vs 36; p=0.0014), with no evidence for a difference after that (Table 7). There also appeared to be a slight decline in S-antibody levels for BNT162b2 vaccinees at later time points (56+ days), and conversely a sustained increase in titres following ChAdOx1 over the same period. Both groups achieved median titres of ≥250U/ml from 7 days after a second dose. Amongst 65-79 year-olds, statistically significant differences in antibody levels were observed between vaccines at ≥28 days (BNT162b2: median 41.4U/ml [IQR 17.8, 84.8] vs ChAdOx1: 30.21U/ml [11.9, 67.16], p<0.0001), and at ≥42 days (BNT162b2: 40.5U/ml [18.64, 81.87] vs ChAdOx1: 35.9U/ml [15, 73.36], p=0.0004) following the first dose (Table 7). However, beyond 56 days, there was insufficient evidence to suggest a difference (BNT162b2: 39.39U/ml [18.9, 75.6] vs ChAdOx1: 36.79U/ml [16.44, 73], p=0.0494).

**Table 7.** Differences in S-antibody levels (U\ml) between demographic and clinical groups, following a single vaccine dose, with vaccine type and clinical categories restricted to the 65-79 years age group. P-values obtained using the Mann-Whitney U test for 2 groups and the Kruskal-Wallis test for >2 groups.

| Table 7 | S-antibody level |  |  | p-value |
| --- | --- | --- | --- | --- |
|  | N | Median | IQR | (* *p* <0.0017) |
| Age groups at 28+ days (dose 1) |  |  |  |  |
| 18-34y | 158 | 59.595 | ( 28.44 , 105 ) | 0.0001* |
| 35-49y | 538 | 37.4 | ( 18.4 , 85.2 ) |  |
| 50-64y | 3,123 | 37.13 | ( 16.28 , 83.6 ) |  |
| 65-79y | 5,631 | 35.89 | ( 14.8 , 75.8 ) |  |
| 80+y | 174 | 25.195 | ( 8.32 , 69.3 ) |  |
| Age 65-79 years |  |  |  |  |
| Vaccine type at 21-27 days (dose 1) |  |  |  |  |
| BNT1612b2 | 229 | 37.71 | ( 13.05 , 96.6 ) | <0.0001* |
| ChAdOx1 | 545 | 13.7 | ( 4.45 , 36.6 ) |  |
| Vaccine type at 28-34d days (dose 1) |  |  |  |  |
| BNT1612b2 | 439 | 44.9 | ( 15.7 , 92.2 ) | <0.0001* |
| ChAdOx1 | 573 | 21.2 | ( 7.94 , 53.7 ) |  |
| Vaccine type at 35-41 days (dose 1) |  |  |  |  |
| BNT1612b2 | 316 | 41.95 | ( 17.09 , 87.4 ) | <0.0001* |
| ChAdOx1 | 423 | 23.4 | ( 8.77 , 58.3 ) |  |
| Vaccine type at 42-55 days (dose 1) |  |  |  |  |
| BNT1612b2 | 577 | 42.39 | ( 19.3 , 90.1 ) | 0.0014* |
| ChAdOx1 | 835 | 36 | ( 13.8 , 73.6 ) |  |
| Vaccine type at 56-69 days (dose 1) |  |  |  |  |
| BNT1612b2 | 790 | 38.855 | ( 19.14 , 74.9 ) | 0.3945 |
| ChAdOx1 | 1047 | 37.2 | ( 17.5 , 75.2 ) |  |
| Vaccine type at 70-84 days (dose 1) |  |  |  |  |
| BNT1612b2 | 275 | 35.3 | ( 14.5 , 64 ) | 0.0637 |
| ChAdOx1 | 338 | 43.96 | ( 16.02 , 84.2 ) |  |
| **28+ days (dose 1) and age 65-79 years** |  |  |  |  |
| **Sex** |  |  |  |  |
| Male | 2585 | 26.5 | ( 10.77 , 60 ) | <0.0001* |
| Female | 3042 | 44.01 | ( 19.6 , 89.7 ) |  |
| **Ethnicity** |  |  |  |  |
| White | 5541 | 35.6 | ( 14.69 , 75.2 ) | 0.0006* |
| South Asian | 38 | 64.295 | ( 32.46 , 192 ) |  |
| Other Asian | 18 | 66.515 | ( 30.12 , 103 ) |  |
| Black | 3 | 46.1 | ( 19.25 , 143 ) |  |
| Mixed | 15 | 60.7 | ( 29.6 , 223 ) |  |
| **Any immunosuppressive therapy** |  |  |  |  |
| No | 4744 | 36.775 | ( 15.6 , 77.1 ) | <0.0001* |
| Yes | 887 | 28.7 | ( 10.44 , 70.6 ) |  |
| **Immunosuppressive therapy following organ transplant** |  |  |  |  |
| No | 5618 | 35.9 | ( 14.86 , 75.8 ) | 0.0157 |
| Yes | 13 | 0.4 | ( 0.4 , 120 ) |  |
| **Immunosuppressive cancer therapy** |  |  |  |  |
| No | 5574 | 35.905 | ( 14.9 , 75.8 ) | 0.0186 |
| Yes | 57 | 17.1 | ( 3.52 , 75.8 ) |  |
| **Steroid tablets** |  |  |  |  |
| No | 5518 | 36.065 | ( 15.04 , 76 ) | <0.0001* |
| Yes | 113 | 16.8 | ( 2.04 , 70.3 ) |  |
| **Steroid inhaler** |  |  |  |  |
| No | 5060 | 36.1 | ( 15.1 , 76.7 ) | 0.0614 |
| Yes | 571 | 31.7 | ( 12.3 , 72.8 ) |  |
| **Non-steroidal immunosuppressive medication** |  |  |  |  |
| No | 5461 | 36.01 | ( 14.98 , 76.4 ) | 0.0002* |
| Yes | 170 | 25.45 | ( 6.21 , 55.6 ) |  |
| **Any cancer diagnosis** |  |  |  |  |
| No | 4877 | 36.5 | ( 15.13 , 76.9 ) | 0.0012* |
| Yes | 754 | 31.385 | ( 11.9 , 65.9 ) |  |
| **Cancer type** |  |  |  |  |
| Haematological cancer | 70 | 7.935 | ( 0.4 , 53 ) | <0.0001* |
| Solid-organ cancer | 684 | 32.5 | ( 13.65 , 67.1 ) |  |
| **Obesity** |  |  |  |  |
| No | 4375 | 37.42 | ( 16.2 , 78 ) | <0.0001* |
| Yes | 1021 | 27.2 | ( 10.35 , 70.3 ) |  |
| **Statin therapy** |  |  |  |  |
| No | 3487 | 39.39 | ( 16.8 , 83.5 ) | <0.0001* |
| Yes | 2144 | 30.025 | ( 11.89 , 64 ) |  |
| **Chronic condition** |  |  |  |  |
| Any chronic condition | 4340 | 33.8 | ( 13.77 , 72.2 ) | <0.0001* |
| No chronic condition | 1291 | 43.83 | ( 18.7 , 88.3 ) |  |
| **Respiratory condition** |  |  |  |  |
| No | 4650 | 36.4 | ( 15.3 , 78.1 ) | 0.0042 |
| Yes | 981 | 32.2 | ( 12.6 , 70.4 ) |  |
| **Diabetes** |  |  |  |  |
| No | 5090 | 36.9 | ( 15.59 , 78.2 ) | <0.0001* |
| Yes | 541 | 22.46 | ( 8.66 , 56 ) |  |
| **Cardiovascular disease** |  |  |  |  |
| No | 3162 | 37.9 | ( 16 , 79 ) | <0.0001* |
| Yes | 2469 | 32.9 | ( 13.5 , 71.3 ) |  |
| **Hypothyroidism** |  |  |  |  |
| No | 5163 | 35.4 | ( 14.51 , 75.6 ) | 0.0371 |
| Yes | 468 | 39.315 | ( 16.76 , 80.1 ) |  |
| **Chronic kidney disease** |  |  |  |  |
| No | 5534 | 36 | ( 15 , 76 ) | 0.0134 |
| Yes | 97 | 21.7 | ( 7.07 , 69.9 ) |  |
| **Liver disease** |  |  |  |  |
| No | 5531 | 35.8 | ( 14.8 , 75.8 ) | 0.7282 |
| Yes | 100 | 40.16 | ( 12.66 , 92.7 ) |  |
| **Neurological condition** |  |  |  |  |
| No | 5519 | 36.01 | ( 14.9 , 76.1 ) | 0.0336 |
| Yes | 112 | 28.525 | ( 8.44 , 63.2 ) |  |
| **28+ days (dose 1) and age 35-49 years** |  |  |  |  |
| **HIV** |  |  |  |  |
| No | 528 | 37.42 | ( 18.54 , 85.3 ) | 0.4742 |
| Yes | 10 | 22.5 | ( 10.89 , 68.5 ) |  |

S-antibody levels appeared higher for females than males and higher following BNT162b2 than ChAdOx1 at ≥28 days after the first dose (all ages) (Table 4). At 14 days after the second dose, all groups had reached average S-antibody levels of 250U/ml or more. Amongst 65-79 year-olds, there was evidence for higher titres in females compared to males (F: median 44.01 U/ml [IQR 19.6, 89.7]) vs M: 26.5 [10.77, 60], p<0.0001) at ≥28 days after the first dose.

There was evidence for a true difference in S-antibody levels by age group at ≥28 days after a single dose (p=0.0001), with lower S-antibody levels seen in older age groups (18-34y: 59.6 U/ml vs ≥80y: 25.2 U/ml) (Table 7). When split by vaccine type, the gradient across the age groups was more apparent amongst BNT162b2 than ChAdOx1 vaccinees (Figure 2, Table 5). All age groups reached median titres of ≥250U/ml from 14 days after a second dose.

Those from minority ethnic groups appeared to have higher titres than those of White ethnicity (p=0.0005 for a difference by ethnicity), although numbers were very small in some groups (Table 7).

At ≥28 days after a single vaccine dose, 65-79 year-olds reporting one or more chronic conditions had lower S-antibody levels than those with no chronic conditions (33.8 U/ml vs 43.83 U/ml; p<0.0001) (Table 7). There was also evidence for lower antibody levels in those with diabetes (22.46 vs 36.9; p<0.0001); those with obesity (27.2 vs 37.42; p<0.0001); those on statin therapy (30.03 vs 39.39; p<0.0001); and those on current immunosuppressive therapy (28.7 vs 36.78; p<0.0001), specifically oral steroids (16.8 vs 36.07; p<0.0001) as well as non-steroidal immunosuppressants (25.45 vs 36.01; p=0.0002). The most marked difference was seen between those with a diagnosis of haematological rather than solid-organ cancer (7.4 vs 31.68; p<0.0001). Additionally there was evidence for a difference in antibody levels by cardiovascular disease status (CVD: 32.9 vs no CVD: 37.9; p<0.0001) and cancer history (cancer: 31.39 vs no cancer: 36.5, p=0.0012), however the absolute differences in levels were small. Though the above disparities were statistically significant, it is not clear whether the absolute differences in antibody levels are of clinical significance with regards to protective immunity. There was insufficient evidence for differences by respiratory condition, hypothyroidism, chronic kidney disease, liver disease, neurological condition, or inhaled steroid therapy status amongst 65-79 year-olds. There was a marked trend towards lower antibody levels amongst 65-79 year-olds receiving immunosuppressive cancer therapy (17.1 vs 35.91; p=0.0186) or immunosuppression following organ transplantation (0.4 vs 35.9; p=0.0157); and in 35-49 year-olds with HIV (22.5 vs 37.42; p=0.4742), but with insufficient evidence to support a true difference. S-antibody levels reached ≥250 U/ml for all clinical groups (including all ages) at ≥14 days after the second dose (Figure 3, Table 6).

## Discussion

Findings from this large community cohort without evidence of prior infection, indicate that the vast majority of individuals have seroconverted to Spike by 4 weeks following the first dose of either BNT162b2 or ChAdOx1 vaccines. While seroconversion following Pfizer/BioNTech’s vaccine appears to occur earlier, and Spike-antibody levels rise earlier compared with Oxford/AstraZeneca’s vaccine, seropositivity rates are equivalent from 4 weeks onwards, and S-antibody levels are comparable from 8 weeks onwards. Following the second dose of either vaccine, seropositivity rises further and S-antibody levels reach ≥250 U/ml for the majority of individuals. Disparities in seroconversion rates and S-antibody levels are evident between demographic and clinical groups after the first dose, particularly amongst those with diagnoses or therapies affecting the immune system. Importantly, S-antibody levels are significantly lower in individuals with common conditions such as diabetes, cardiovascular disease, and obesity.

A study of a large healthcare worker cohort found that seroconversion rates were 99.5% (2706/2720) and 97.1% (864/890) at >14 days after a single dose of BNT162b2 and ChAdOx1, respectively^23^. The slightly lower seroconversion rates observed in our cohort could be attributed to the older average age and higher prevalence of comorbidities, and thus a reduced healthy-worker bias. Conversely, seroconversion rates appear substantially higher in our cohort than those reported by the REACT2 study^35^ which found 84.1% (82.2, 85.9) seroconversion ≥21 days after a dose of BNT162b2; though this may be attributable to differences in assay sensitivities.

A large observational study including 38,262 SARS-CoV-2-naive individuals (from a community cohort enrolled in the UK’s national COVID-19 Infection Survey), modelled the probability of seroconversion to Spike following a single dose vaccination^27^. Similar to our findings, they reported lower probabilities of seroconversion, as well as slower rate of increase of S-antibody titres, following ChAdOx1 compared with BNT162b2; also in line with our findings, differences between the two vaccines attenuated at later time points following the first dose. For BNT1612b2, the authors reported a gradual decline in anti-S levels from 35 days post-first dose, whereas titres remained stable for ChAdOx1. We also observed a very slight decline over this time period for BNT162b2 but noted a slight and sustained increase in titres for ChAdOx1 over the same time period. It is not possible to compare the magnitude of these effects across the studies due to lack of standardisation across the different assays; nor is it yet clear whether these trends are of clinical significance.

Both trial and observational data relating to single-dose BNT162b2^36–39^ and ChAdOx1^27,40^ indicate lower antibody titres in older adults compared to younger adults, which is also reflected in our findings. While evidence directly linking S-antibody titres with efficacy across different age groups is lacking, available data do suggest lower VE against infection in older populations^11,41^. However, VE against clinical outcomes appears similar across age groups^9,42^; it is possible that differences are masked by the uncertainty in the available VE estimates, or it may be that protective levels of antibodies are achieved once a certain threshold is passed.

Notably, those with haematological malignancy and those on immunosuppressive medications, especially following organ transplantation, showed markedly lower seroconversion rates after the first vaccine dose. This is in line with findings from studies focusing on patients with cancer^43^, and specifically those with haematological malignancy^44^, as well as immunosuppression^45,46^, particularly organ transplant recipients^47–49^. Taken together, these findings highlight the need to further investigate vaccine immunogenicity and efficacy in these groups. Early data hinting at the correlation of S-antibody levels with protection against infection^50^ suggest that the immunological and clinical impact of expediting the second dose of vaccination in these groups should be explored.

Our findings provide immunological insights to support real-world data on vaccine effectiveness (VE). Reports of single-dose VE from the UK indicate lower odds of infection from 21 days after a single dose of either vaccine (ChAdOx1 - OR 0.36 [95% CI 0.30, 0.45]; BNT162b2 0.33 [0.28, 0.39]) in a large community cohort^11^. Data on BNT162b2 from Israel indicate onset of protection from as early as 14 days after the first dose, against asymptomatic (14-20d: 29% [95%CI 17,39]; 21-27d: 52% [41,60]) and symptomatic (57% [50,63]; 66% [57,73]) infections^42^. Several studies also report reduced PCR Cycle threshold values, indicative of viral load and potential infectivity^51,52^, from as early as 12 days following a single dose^11,13,15^. Across most reports, protective effects first appear between 2-4 weeks after a single dose depending on the outcome studied, with slightly longer taken to reach peak VE with ChAdOx1 than BNT162b2^12^; these trends mirror the immunogenicity data on seroconversion rates and S-antibody titres reported here and elsewhere^27^. There is also emerging evidence from other studies to suggest lower VE in those with diabetes and cardiovascular conditions^41^.

Accumulating evidence suggests the importance of T-cell responses in protection against COVID-19^50,53,54^, and it may be that, at earlier time points when S-antibody levels following ChAdOx1 are lower than BNT162b2, VE is driven by T-cell immunity. Furthermore, in those with immunosuppression or haematological malignancy, it is possible that helper T-cell impairment contributes to attenuated antibody responses. As has been noted with immune responses to infection^55^, it is possible that the overall balance and co-ordination between humoral and cellular arms of the adaptive immune response may be the best predictor of protection^56^, rather than the level of any one immune component alone. In future work, it will be important to link data from deeper immunophenotyping with the risk of vaccine breakthroughs and failures, and their clinical severity and viral load, in order to better understand immunity against these outcomes.

### Strengths and Limitations

The strengths of this study include the large community cohort, which spans many ages and ethnic groups, and includes large numbers of individuals with common chronic conditions as well as capturing those with rarer diagnoses. These data constitute some of the earliest real-world evidence on immunogenicity for many clinical groups, and for ChAdOx1 more broadly, and are likely to be generalisable to other high-income settings deploying these vaccines. We employed a highly sensitive, widely used validated commercial assay that gives quantitative readouts on a linear scale, allowing these data to be easily understood, replicated, and informative for clinical practitioners. Limitations include the use of self-reported vaccination status, which may have introduced error into the assignment of vaccination date and type, or the omission of a recent second dose for some individuals; and collection of clinical data at enrolment only, which may have led to misclassification due to more recent diagnoses. Additionally, the cancer categories encompassed both prior and current diagnoses. Due to the available assay platform and dilution capabilities, the dynamic range of the S-antibody assay was limited to 0.4U/ml - 250U/ml, which precluded differentiation of S-antibody levels between groups following the second dose and amongst those with prior infection. Due to small numbers of individuals with HIV, immunosuppressive cancer therapy, and immunosuppression following organ transplantation, there was likely insufficient power to demonstrate a difference in S-antibody levels. With regards to immunosuppressive therapies such as steroids, attenuation of vaccine responses is likely to be dose-dependent, however we were not able to ascertain dosage or duration of therapy; and we were also unable to distinguish specific drugs such as Methotrexate or Rituximab, which are most likely to affect humoral responses^21,44,46,57^.

### Conclusions

Our data indicate very high rates of seroconversion to Spike following a single dose of either vaccine, with near-complete seroconversion following a second dose, in individuals with no evidence of prior infection. Despite earlier responses with BNT162b2 compared to ChAdOx1, both vaccines demonstrate equivalent seropositivity rates and S-antibody levels from 4 and 8 weeks following a single dose, respectively, with no differences in seropositivity seen after a second dose. Disparities in seropositivity rates between demographic and clinical groups also do not persist after the second dose. High seroconversion rates after the first dose lend support to the UK policy to prioritise first-dose coverage across the population, however our data also suggest attenuated immune responses in some clinical groups, which warrant further investigation. Studying longer-term dynamics of the humoral and cellular immune responses, and the correlates of protection against disease, asymptomatic infection, and onward transmission, including for emerging variants of concern, remain key questions for informing global COVID-19 control policies.

## Data Availability

We aim to share aggregate data from this project on our website and via a “Findings so far” section on our website - https://ucl-virus-watch.net/. We will also be sharing individual record level data on a research data sharing service such as the Office of National Statistics Secure Research Service. In sharing the data we will work within the principles set out in the UKRI Guidance on best practice in the management of research data. Access to use of the data whilst research is being conducted will be managed by the Chief Investigators (ACH and RWA) in accordance with the principles set out in the UKRI guidance on best practice in the management of research data. We will put analysis code on publicly available repositories to enable their reuse.

## Data availability

We aim to share aggregate data from this project on our website and via a “Findings so far” section on our website - https://ucl-virus-watch.net/. We will also be sharing individual record level data on a research data sharing service such as the Office of National Statistics Secure Research Service. In sharing the data, we will work within the principles set out in the UKRI Guidance on best practice in the management of research data. Access to use of the data whilst research is being conducted will be managed by the Chief Investigators (ACH and RWA) in accordance with the principles set out in the UKRI guidance on best practice in the management of research data. We will put analysis code on publicly available repositories to enable their reuse.

## References

1. World Health Organization. Coronavirus (COVID-19) Dashboard. Available from: https://covid19.who.int/. (accessed 11 May 2021).

2. World Health Organization. Draft Landscape of COVID-19 Candidate Vaccines. Available from: https://www.who.int/publications/m/item/draft-landscape-of-covid-19-candidate-vaccines. (accessed 11 May 2021).

3. Medicines and Healthcare Products Regulatory Agency. Regulatory approval of Pfizer-BioNTech vaccine for COVID-19. Available from: https://www.gov.uk/government/publications/regulatory-approval-of-pfizer-biontech-vaccine-for-covid-19. (accessed 11 May 2021).

4. Medicines and Healthcare Products Regulatory Agency. Regulatory approval of AstraZeneca vaccine for COVID-19. Available from: https://www.gov.uk/government/publications/regulatory-approval-of-covid-19-vaccine-astrazeneca. (accessed 11 May 2021).

5. Ju, B. et al. Human neutralizing antibodies elicited by SARS-CoV-2 infection. Nature 584, 115–119 (2020).

6. Oliveira, S. C., de Magalhães, M. T. Q. & Homan, E. J. Immunoinformatic Analysis of SARS-CoV-2 Nucleocapsid Protein and Identification of COVID-19 Vaccine Targets. Front. Immunol. 11, 1–10 (2020).

7. Department of Health and Social Care. Priority groups for coronavirus (COVID-19) vaccination - advice from the JCVI - 30 December 2020. Available from: https://www.gov.uk/government/publications/priority-groups-for-coronavirus-covid-19-vaccination-advice-from-the-jcvi-30-december-2020. (accessed 11 May 2021).

8. Public Health England. Coronavirus (COVID-19) in the UK - Vaccinations. Available from: https://coronavirus.data.gov.uk/details/vaccinations. (accessed 11 May 2021).

9. Polack, F. P. et al. Safety and Efficacy of the BNT162b2 mRNA Covid-19 Vaccine. N. Engl. J. Med. 383, 2603–2615 (2020).

10. Voysey, M. et al. Safety and efficacy of the ChAdOx1 nCoV-19 vaccine (AZD1222) against SARS-CoV-2: an interim analysis of four randomised controlled trials in Brazil, South Africa, and the UK. Lancet 397, 99–111 (2021).

11. Pritchard, E. et al. Impact of vaccination on SARS-CoV-2 cases in the community: a population-based study using the UK’s COVID-19 Infection Survey. medRxiv (2021).

12. Bernal, JL, et al. Effectiveness of the Pfizer-BioNTech and Oxford-AstraZeneca vaccines on covid-19 related symptoms, hospital admissions, and mortality in older adults in England: test negative case-control study. bmj 373 (2021).

13. Levine-Tiefenbrun, M. et al. Initial report of decreased SARS-CoV-2 viral load after inoculation with the BNT162b2 vaccine. Nat. Med. (2021).

14. Shah, A. et al. Effect of vaccination on transmission of COVID-19: an observational study in healthcare workers and their households. medRxiv (2021).

15. Harris, R., et al. Impact of vaccination on household transmission of SARS-COV-2 in England. 2021. Available from: https://khub.net/documents/135939561/390853656/Impact+of+vaccination+on+household+transmission+of+SARS-COV-2+in+England.pdf/35bf4bb1-6ade-d3eb-a39e-9c9b25a8122a?t=1619601878136. (Accessed 19 May 2021).

16. Department of Health and Social Care. Optimising the COVID-19 vaccination programme for maximum short-term impact. Available from: https://www.gov.uk/government/publications/prioritising-the-first-covid-19-vaccine-dose-jcvi-statement/optimising-the-covid-19-vaccination-programme-for-maximum-short-term-impact. (Accessed 11 May 2021).

17. Docherty, A. B. et al. Features of 20 133 UK patients in hospital with covid-19 using the ISARIC WHO Clinical Characterisation Protocol: prospective observational cohort study. BMJ m1985 (2020).

18. Yates, T. et al. Obesity, ethnicity and risk of critical care, mechanical ventilation and mortality in patients admitted to hospital with COVID□19: Analysis of the ISARIC CCP□UK cohort. Obesity oby. 23178 (2021).

19. McGurnaghan, S. J. et al. Risks of and risk factors for COVID-19 disease in people with diabetes: a cohort study of the total population of Scotland. Lancet Diabetes Endocrinol. 9, 82–93 (2021).

20. Mathur, R. et al. Ethnic differences in SARS-CoV-2 infection and COVID-19-related hospitalisation, intensive care unit admission, and death in 17 million adults in England: an observational cohort study using the OpenSAFELY platform. Lancet 397, 1711–1724 (2021).

21. Arnold, J., Winthrop, K. & Emery, P. COVID-19 vaccination and antirheumatic therapy. Rheumatology 1–7 (2021).

22. Angyal, A. et al. T-cell and antibody responses to first BNT162b2 vaccine dose in previously SARS-CoV-2-infected and infection-naive UK healthcare workers: a multicentre, prospective, observational cohort study. SSRN Electron. J. (2021).

23. Eyre, D. W. et al. Quantitative SARS-CoV-2 anti-spike responses to Pfizer-BioNTech and Oxford-AstraZeneca vaccines by previous infection status. medRxiv (2021).

24. Tut, G. et al. Profile of Humoral and Cellular Immune Responses to Single BNT162b2 or ChAdOx1 Vaccine in Residents and Staff Within Residential Care Homes (VIVALDI Study). SSRN Electron. J. (2021).

25. Abu Jabal, K. et al. Impact of age, ethnicity, sex and prior infection status on immunogenicity following a single dose of the BNT162b2 mRNA COVID-19 vaccine: real-world evidence from healthcare workers, Israel, December 2020 to January 2021. Eurosurveillance 26, 1–5 (2021).

26. Subbarao, S. et al. Robust antibody responses in 70 – 80-year-olds 3 weeks after the first or second doses of Pfizer / BioNTech COVID-19 vaccine, United Kingdom, January to February 2021. Eurosurveillance 26, 1–6 (2021).

27. Wei, J. et al. The impact of SARS-CoV-2 vaccines on antibody responses in the general population in the United Kingdom. medRxiv (2021)

28. Hayward, A. et al. Risk factors, symptom reporting, healthcare-seeking behaviour and adherence to public health guidance: Protocol for Virus Watch, a prospective community cohort study. medRxiv (2020).

29. Muench, P. et al. Development and Validation of the Elecsys Anti-SARS-CoV-2 Immunoassay as a Highly Specific Tool for Determining Past Exposure to SARS-CoV-2. J. Clin. Microbiol. 58, (2020).

30. Ainsworth, M. et al. Performance characteristics of five immunoassays for SARS-CoV-2: a head-to-head benchmark comparison. Lancet Infect. Dis. 20, 1390–1400 (2020).

31. Public Health England. Evaluation of Roche Elecsys Anti-SARS-CoV-2 serology assay for the detection of anti-SARS-CoV-2 antibodies. Available from: <https://assets.publishing.service.gov.uk/government/uploads/system/uploads/attachment_data/file/891598/Evaluation_of_Roche_Elecsys_anti_SARS_CoV_2_PHE_200610_v8.1_FINAL.pdf>. (Accessed 11 May 2021).

32. Poljak, M., Oštrbenk Valenčak, A., štamol, T. & Seme, K. Head-to-head comparison of two rapid high-throughput automated electrochemiluminescence immunoassays targeting total antibodies to the SARS-CoV-2 nucleoprotein and spike protein receptor binding domain. J. Clin. Virol. 137, (2021).

33. Riester, E. et al. Performance evaluation of the Roche Elecsys Anti-SARS-CoV-2 S immunoassay. medRxiv (2021).

34. Roche Diagnostics. Press release: Roche launches new quantitative antibody test to measure SARS-CoV-2 antibodies, to support the evaluation of vaccines. Available from: https://www.roche.com/media/releases/med-cor-2020-09-18b.htm. (Accessed 11 May 2021).

35. Ward, H. et al. REACT-2 Round 5: increasing prevalence of SARS-CoV-2 antibodies demonstrate impact of the second wave and of vaccine roll-out in England. medRxiv (2021).

36. Walsh, E. E. et al. Safety and Immunogenicity of Two RNA-Based Covid-19 Vaccine Candidates. N. Engl. J. Med. 383, 2439–2450 (2020).

37. Kontopoulou, K. et al. Immunogenicity after the First Dose of the BNT162b2 mRNA COVID-19 Vaccine: Real-World Evidence from Greek Healthcare Workers. SSRN Electron. J. (2021).

38. Müller, L. et al. Age-dependent immune response to the Biontech/Pfizer BNT162b2 COVID-19 vaccination. Clin. Infect. Dis. (2021) doi:10.1093/cid/ciab381.

39. Collier, D. A. et al. Age-Related Heterogeneity in Neutralising Antibody Responses to SARS-CoV-2 Following BNT162b2 Vaccination. SSRN Electron. J. (2021).

40. Ramasamy, M. N. et al. Safety and immunogenicity of ChAdOx1 nCoV-19 vaccine administered in a prime-boost regimen in young and old adults (COV002): a single-blind, randomised, controlled, phase 2/3 trial. Lancet 396, 1979–1993 (2020).

41. Yelin, I. et al. Associations of the BNT162b2 COVID-19 vaccine effectiveness with patient age and comorbidities. medRxiv (2021).

42. Dagan, N. et al. BNT162b2 mRNA Covid-19 Vaccine in a Nationwide Mass Vaccination Setting. N. Engl. J. Med. 1–12 (2021)..

43. Monin, L. et al. Safety and immunogenicity of one versus two doses of the COVID-19 vaccine BNT162b2 for patients with cancer: interim analysis of a prospective observational study. Lancet. Oncol. 1–14 (2021).

44. Herishanu, Y. et al. Efficacy of the BNT162b2 mRNA COVID-19 Vaccine in Patients with Chronic Lymphocytic Leukemia. Blood (2021).

45. Geisen, U. M. et al. Immunogenicity and safety of anti-SARS-CoV-2 mRNA vaccines in patients with chronic inflammatory conditions and immunosuppressive therapy in a monocentric cohort. Ann. Rheum. Dis. 1–6 (2021).

46. Deepak, P. et al. Glucocorticoids and B Cell Depleting Agents Substantially Impair Immunogenicity of mRNA Vaccines to SARS-CoV-2. medRxiv (2021).

47. Boyarsky, B. J. et al. Immunogenicity of a Single Dose of SARS-CoV-2 Messenger RNA Vaccine in Solid Organ Transplant Recipients. JAMA - J. Am. Med. Assoc. 2–4 (2021).

48. Rabinowich, L. et al. Low immunogenicity to SARS-CoV-2 vaccination among liver transplant recipients. J. Hepatol. (2021).

49. Marinaki, S. et al. Immunogenicity of SARS□CoV□2 BNT162b2 vaccine in solid organ transplant recipients. Am. J. Transplant. ajt. 16607 (2021).

50. Wyllie, D. et al. SARS-CoV-2 responsive T cell numbers and anti-Spike IgG levels are both associated with protection from COVID-19: A prospective cohort study in keyworkers. medRxiv (2021).

51. Singanayagam, A. et al. Duration of infectiousness and correlation with RT-PCR cycle threshold values in cases of COVID-19, England, January to May 2020. Eurosurveillance 25, (2020).

52. Lennard, Y. L. et al. SARS-CoV-2 infectivity by viral load, S gene variants and demographic factors and the utility of lateral flow devices to prevent transmission. medRxiv (2021).

53. Kalimuddin, S. et al. Early T cell and binding antibody responses are associated with COVID-19 RNA vaccine efficacy onset. Med 1–7 (2021).

54. McMahan, K. et al. Correlates of protection against SARS-CoV-2 in rhesus macaques. Nature 590, 630–634 (2021).

55. Rydyznski Moderbacher, C. et al. Antigen-Specific Adaptive Immunity to SARS-CoV-2 in Acute COVID-19 and Associations with Age and Disease Severity. Cell 183, 996–1012.e19 (2020).

56. Poland, G. A., Ovsyannikova, I. G. & Kennedy, R. B. SARS-CoV-2 immunity: review and applications to phase 3 vaccine candidates. Lancet 396, 1595–1606 (2020).

57. Baker, D. et al. COVID-19 vaccine-readiness for anti-CD20-depleting therapy in autoimmune diseases. Clin. Exp. Immunol. 202, 149–161 (2020).

