## Supplementary material for "Spike-antibody responses to ChAdOx1 and BNT162b2 vaccines by demographic and clinical factors (Virus Watch study)": (Table S1)

**Supplementary Materials:**

**Table S1** - Demographic features of Virus Watch study cohort and sub-cohort undergoing monthly antibody testing, compared with Office for National Statistics (UK) data for England and Wales.

| **Characteristic** | **Monthly Antibody Testing Sub-Cohort** | **All Virus Watch participants**  (3rd May 2021) | **ONS England & Wales (%)*** |
| --- | --- | --- | --- |
| **Total** | 11,102 | **50,648** | **100%** |
| **Age groups** |  |  |  |
| <24y | 186 (1.68) | 9700 (19.3) | 29.70% |
| 25-44y | 1,389 (12.51) | 10,500 (21%) | 26.10% |
| 45-64y | 4,412 (39.74) | 16,635 (33%) | 25.60% |
| 65+y | 5115 (46.07) | 13,813 (27%) | 18.50% |
| **Ethnicity** |  |  |  |
| White | 10,711 (96.48) | 37310 (73.2%) | 85.80% |
| Mixed | 89 (0.8) | 890 (1.8%) | 2.20% |
| South Asian | 154 (1.39) | 2,639 (5.2%) | 5.30% |
| Other Asian | 64 (0.58) | 399 (0.8%) | 2.20% |
| Black | 35 (0.32) | 458 (0.9%) | 3.30% |
| Other/PNTS**/Missing | 49 (0.44) | 8952 (17.8) | 1% |
| **Sex** |  |  |  |
| Male | 4,766 (42.93) | 18,849 (37%) | 49.40% |
| Female | 6,321 (56.94) | 23,378 (46%) | 50.60% |
| Other/PNTS**/Missing | 15 (0.14) | 8,421 (17%) |  |
| **Region** |  |  |  |
| East Midlands | 961 (8.66) | 4,169 (8.2%) | 4.50% |
| East of England | 2625 (23.64) | 9,421 (19%) | 12.40% |
| London | 1237 (11.14) | 8,384 (17%) | 9.30% |
| North East | 570 (5.13) | 2,221 (4.4%) | 8.10% |
| North West | 1215 (10.94) | 4,668 (9.2%) | 10% |
| South East | 2253 (20.29) | 8,333 (16%) | 10.50% |
| South West | 822 (7.4) | 3,151 (6.2%) | 15.10% |
| Wales | 216 (1.95) | 1,050 (2.1%) | 15.40% |
| West Midlands | 570 (5.13) | 2,352 (4.6%) | 9.50% |
| Yorkshire and The Humber | 607 (5.47) | 2,469 (4.9%) | 5.30% |
| Other/Missing | 26 (0.24) |  |  |
| **No. of householders*** |  |  |  |
| 1 | 2615 (23.55) | 7,727 (15%) | 29.50% |
| 2 | 6733 (60.65) | 21,761 (43%) | 34.50% |
| 3 | 820 (7.39) | 7,627 (15%) | 15.40% |
| 4 | 696 (6.27) | 8,823 (17%) | 13.90% |
| 5 | 185 (1.67) | 3,353 (6.6%) | 4.50% |
| 6 | 38 (0.34) | 1,357 (2.7%) | 2.10% |
| *ONS household figures from:  https://www.ethnicity-facts-figures.service.gov.uk/housing/housing-conditions/overcrowded-households/latest | | | |
| **Prefer not to say |  |  |  |

**Table S2** - Survey questions used to collect self-reported data included in this analysis.

| **Collection** | **Question** | **Response options** | **Analysis variables** |
| --- | --- | --- | --- |
| Enrollment | Date of birth | DD/MM/YYYY | **Age** |
|  | Could you please confirm each person’s sex at birth? | 1. Male  2. Female  3. Intersex  4. Prefer not to say | **Sex:**  Male (1)  Female (2)  Other / Prefer not to say (3,4) |
|  | What is your ethnic group? | 1. White - English/ Welsh/ Scottish/ Northern Irish/ British  2. White – Irish  3. White - Gypsy or Irish Traveller  4. Any other white background (please describe)  5. Asian/ Asian British - Indian Asian/ Asian British – Pakistani  6. Asian/ Asian British – Bangladeshi  7. Asian/ Asian British – Chinese  8. Any other Asian/ Asian British background (please describe)  9. Black African  10. Black Caribbean  11. Any other Black  12. African/ Caribbean background (please describe)  13. Arab  14. Any other ethnic group (please describe)  15. Mixed/ multiple ethnic groups - White and Black Caribbean  16. Mixed/ multiple ethnic groups - White and Black African  17. Mixed/ multiple ethnic groups - White and Asian  18. Any other mixed/ multiple ethnic background (please describe)  19. Prefer not to say | **Ethnicity:**  White (1,2,3,4)  South Asian (5,6)  Other Asian (7,8)  Black (9,10,11,12)  Mixed (15, 16, 17, 18)  Other / Prefer not to say (13, 14, 19) |
|  | Has a doctor or other health professional ever told you that you have any of these conditions?  Please select all that apply | 1. Asthma  2. Arthritis  3. Congestive heart failure  4. Coronary heart disease  5. Angina  6. Heart attack or myocardial infarction  7. Stroke  8. Emphysema  9. Chronic bronchitis  10. COPD (Chronic Obstructive Pulmonary Disease)  11. Cystic fibrosis  12. Hypothyroidism or an under-active thyroid  13. Any kind of liver condition  14. Cancer or malignancy  15. Insulin treated diabetes  16. Other diabetes  17. Epilepsy  18. High blood pressure/hypertension  19. An emotional, nervous or psychiatric problem  20. Multiple Sclerosis  21. HIV  22. Chronic kidney disease  23. Conditions affecting the brain and nerves, such as Parkinson's disease, motor neurone disease, multiple sclerosis (MS), a learning disability or cerebral palsy  24. Problems with your spleen or you've had your spleen removed  25. Sickle cell disease  26. Other long standing/chronic condition  27. None of these | **Health conditions:**  No chronic condition (27 and nil else)  Respiratory condition (1,8,9,10,11)  Cardiovascular disease (3,4,5,6,7,18)  Diabetes (15,16)  Hypothyroidism (12)  Chronic kidney disease (22)  Neurological conditions (17,20,23)  Liver condition (13)  HIV (21)    **Cancer:**  Any (14) |
|  | [If selected 14 above] What type of cancer or malignancy was that? Please select all that apply  (MALE) | 1. Bowel/colorectal  2. Lung  3. Breast  4. Prostate  5. Liver  6. Skin cancer or melanoma  7. Blood or bone marrow cancer, such as leukaemia  8. Other | **Cancer:**  Haematological (7)  Non-haematological (1,2,3,4,5,6,8) |
|  | [If selected 14 above] What type of cancer or malignancy was that? Please select all that apply  (FEMALE) | 1. Bowel/colorectal  2. Lung  3. Breast  4. Liver  5. Skin cancer or melanoma  6. Blood or bone marrow cancer, such as leukaemia  7. Other | **Cancer:**  Haematological (6)  Non-haematological (1,2,3,4,5,7) |
|  | Are you currently receiving treatment or taking medications that may affect your immune system? Please select all that apply | 1. Medication following an organ transplant  2. Medicines such as steroid tablets that weaken the immune system  3. Targeted therapy or chemotherapy for cancer treatment  4. Radiotherapy for cancer treatment  5. Other treatment or medication that may affect immune system  6. None of these | **Immunosuppressive therapy:**  Any (1,2,3,4 or Steroids as below)  Post-organ transplant (1)  Cancer therapy (3,4)  Non-steroid (2,5 excluding those on Steroids as below) |
|  | Which of the following medicines do you take? Please select all that apply | 1. Regularly taking Aspirin  2. Regularly taking ""NSAIDS"" e.g. Ibuprofen, nurofen, diclofenic, naproxen.  3. Regularly taking blood pressure medicines ending in ""-pril"" such as enalapril, lisinopril, captopril, ramipril  4. Regularly taking blood pressure measurements ending in ""-sartan"" such as losartan, valsartan, irbesartan  5. Regularly taking anticoagulants e.g warfarin, ivaroxaban (Xarelto), dabigatran (Pradaxa), apixaban (Eliquis), edoxaban (Lixiana)  6. Steroid tablets  7. Regularly use a steroid inhaler  8. Regularly take statins e.g. atorvastatin (Lipitor)  9. None of these | **Immunosuppressive therapy:**  Steroid tablet (6)  Steroid inhaler (7)    **Statins** (8) |
|  | What is your height in centimetres (cm)? Please enter digits only, e.g. '5' and not 'five’  *OR*  How many feet tall are you (rounded down)? Please enter the feet component of your height. For example if you're 5 foot 4, please enter 5  *&*  How many inches tall are you above your feet value? Please enter the inches component of your height. For example if you're 5 foot 4, please enter 4 | | **BMI** (kg/m^2) |
|  | How much do you weigh in kilograms (kg)? Please enter digits only, e.g. '5' and not 'five'  *OR*  What is your weight in stone, rounded down? For example if you are 8 stone, 10 pounds, please enter 8. If you do not use stone, please feel free to leave this blank and enter your weight fully in pounds  *&/OR*  How much do you weight in pounds (lbs) (above your stone weight)? For example, if you are 8 stone, 10 pounds, please enter 10.If you did not enter a value for stone, please enter your weight fully in lbs here | |  |
|  | Have you received a letter from the NHS, saying that "*The NHS has identified you as someone at risk of severe illness if you catch coronavirus, because you have an underlying disease or health condition that means if you catch the virus, you are more likely to be admitted to hospital than others"?* | | **NHS Risk Letter** (Y) |
| January 2021 | Has anyone in the household ever received a COVID-19 vaccine in the past? | Yes  No  Unsure (e.g. as part of a blinded COVID-19 trial) | **Vaccination status** |
| February – May 2021 | Has anyone in the household received a COVID-19 vaccine in the past week? | Yes  No  Unsure (e.g. as part of a blinded COVID-19 trial) | **Vaccination status** |
| January – May 2021 | Please select which dose(s) of the COVID-19 vaccine you received? | 1st Dose  2nd Dose | **Vaccination status** |
|  | What date did you receive the 1st dose? (dd-mm-yyyy) Please provide an estimate if you cannot recall the date | DD-MM-YYYY | **Vaccination date** |
|  | Which type of vaccine did you receive as the 1^st^ / 2^nd^ dose? | 1. Pfizer Biontech vaccine  2. Oxford AstraZeneca vaccine  3. Moderna vaccine  4. Other vaccine  5. Don't know/Don't remember | **Vaccination type:**  BNT162b2 (1)  ChAdOx1 (2) |

**Figure S1 -** Study inclusion flow-chart.

*Number of samples	Samples excluded*

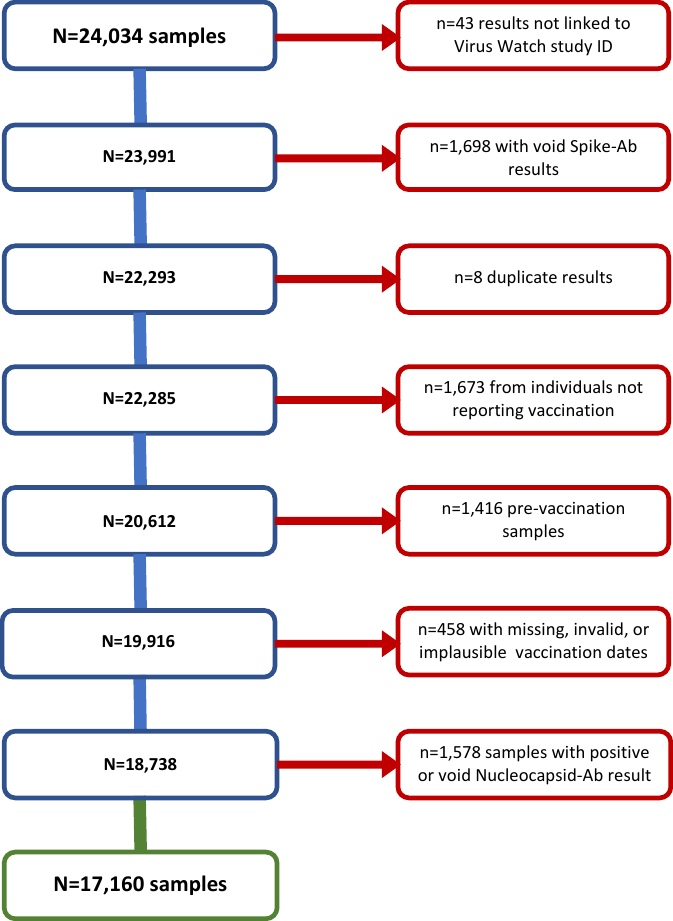

STROBE checklist

|  | Item No | Recommendation | Page No |
| --- | --- | --- | --- |
| **Title and abstract** | | |  |
|  | 1 | (*a*) Indicate the study's design with a commonly used term in the title or the abstract | 1,2 |
|  |  | (*b*) Provide in the abstract an informative and balanced summary of what was done and what was found | 2 |
| **Introduction** | | |  |
| Background/rationale | 2 | Explain the scientific background and rationale for the investigation being reported | 3 |
| Objectives | 3 | State specific objectives, including any prespecified hypotheses | 3 |
| **Methods** | | |  |
| Study design | 4 | Present key elements of study design early in the paper | 3,4,5 |
| Setting | 5 | Describe the setting, locations, and relevant dates, including periods of recruitment, exposure, follow-up, and data collection | 4,5 |
| Participants | 6 | *a) Cohort study*? Give the eligibility criteria, and the sources and methods of selection of participants. Describe methods of follow-up  *Case-control study*? Give the eligibility criteria, and the sources and methods of case ascertainment and control selection. Give the rationale for the choice of cases and controls  *Cross sectional study*? Give the eligibility criteria, and the sources and methods of selection of participants | 4,5,6 |
|  |  | (*b*) *Cohort study*? For matched studies, give matching criteria and number of exposed and unexposed  *Case-control study*? For matched studies, give matching criteria and the number of controls per case | N/A |
| Variables | 7 | Clearly define all outcomes, exposures, predictors, potential confounders, and effect modifiers. Give diagnostic criteria, if applicable | 4,5,6, Supplementary Table S2 |
| Data sources/ measurement | 8* | For each variable of interest, give sources of data and details of methods of assessment (measurement). Describe comparability of assessment methods if there is more than one group | 4,5,6 |
| Bias | 9 | Describe any efforts to address potential sources of bias | 4,5,6 |
| Study size | 10 | Explain how the study size was arrived at | 4, Supplementary Figure S1 |
| Quantitative variables | 11 | Explain how quantitative variables were handled in the analyses. If applicable, describe which groupings were chosen and why | 4,5,6 |
| Statistical methods | 12 | (*a*) Describe all statistical methods, including those used to control for confounding | 5,6 |
|  |  | (*b*) Describe any methods used to examine subgroups and interactions | 5,6 |
|  |  | (*c*) Explain how missing data were addressed | 4,5 |
|  |  | (*d*) *Cohort study*? If applicable, explain how loss to follow-up was addressed  *Case-control study*? If applicable, explain how matching of cases and controls was addressed  *Cross sectional study*? If applicable, describe analytical methods taking account of sampling strategy | N/A |
|  |  | (*e*) Describe any sensitivity analyses | N/A |
| **Results** | | |  |
| Participants | 13* | (*a*) Report numbers of individuals at each stage of study? E.g. numbers potentially eligible, examined for eligibility, confirmed eligible, included in the study, completing follow-up, and analysed. | Supplementary Figure S1 |
|  |  | (*b*) Give reasons for non-participation at each stage | 4,5 Supplementary Figure S1 |
|  |  | (*c*) Consider use of a flow diagram | Supplementary Figure S1 |
| Descriptive data | 14* | (*a*)Give characteristics of study participants (e.g. demographic, clinical, social) and information on exposures and potential confounders | Table 1, 2 |
|  |  | (*b*) Indicate number of participants with missing data for each variable of interest | Table 1, 2 |
|  |  | (*c*) *Cohort study*? Summarise follow-up time (e.g. average and total amount) | 4, Table 2 |
| Outcome data | 15* | *Cohort study*? Report numbers of outcome events or summary measures over time | Table 3,4,5,6 |
|  |  | *Case-control study?* Report numbers in each exposure category, or summary measures of exposure |  |
|  |  | *Cross sectional study?* Report numbers of outcome events or summary measures |  |
| Main results | 16 | (*a*) Report the numbers of individuals at each stage of the study? E.g. numbers potentially eligible, examined for eligibility, confirmed eligible, included in the study, completing follow-up, and analysed. | Supplementary Figure S1 |
|  |  | (*b*) Give reasons for non-participation at each stage | Supplementary Figure S1 |
|  |  | (*c*) Consider use of a flow diagram | Supplementary Figure S1 |
| Other analyses | 17 | Report other analyses done? E.g. analyses of subgroups and interactions, and sensitivity analyses | 7,8,9 |
| **Discussion** | | |  |
| Key results | 18 | Summarise key results with reference to study objectives | 9 |
| Limitations | 19 | Discuss limitations of the study, taking into account sources of potential bias or imprecision. Discuss both direction and magnitude of any potential bias | 11 |
| Interpretation | 20 | Give a cautious overall interpretation of results considering objectives, limitations, multiplicity of analyses, results from similar studies, and other relevant evidence | 11 |
| Generalisability | 21 | Discuss the generalisability (external validity) of the study results | 11 |
| **Other information** | | |  |
| Funding | 22 | Give the source of funding and the role of the funders for the present study and, if applicable, for the original study on which the present article is based | 6,12 |
